# The immunogenetics of viral antigen response is associated with sub-type specific glioma risk and survival

**DOI:** 10.1101/2021.09.13.21263349

**Authors:** Geno Guerra, Linda Kachuri, George Wendt, Helen M. Hansen, Steven J. Mack, Annette M. Molinaro, Terri Rice, Paige Bracci, John K. Wiencke, Nori Kasahara, Jeanette E Eckel-Passow, Robert B. Jenkins, Margaret Wrensch, Stephen S. Francis

## Abstract

Glioma is a highly fatal cancer with prognostically significant molecular subtypes and few known risk factors. Multiple studies have implicated infections in glioma susceptibility, but evidence remains inconsistent. Genetic variants in the human leukocyte antigen (HLA) region modulate host response to infection and have been linked to glioma risk. In this study we leveraged genetic predictors of antibody response to 10 viral antigens to investigate the relationship with glioma risk and survival.

Genetic reactivity scores (GRS) for each antigen were derived from genome-wide significant (p<5×10^−8^) variants associated with immunoglobulin G antibody response in the UK Biobank cohort. We conducted parallel analyses of glioma risk and survival for each GRS and HLA alleles imputed at two-field resolution using data from 3418 glioma patients subtyped by somatic mutations and 8156 controls.

Genetic reactivity scores to Epstein-Barr virus (EBV) ZEBRA and EBNA antigens and Merkel cell polyomavirus (MCV) VP1 antigen were associated with glioma risk and survival (Bonferroni-corrected p<0.01). GRSZEBRA and GRSMCV were associated in opposite directions with risk of *IDH* wild type gliomas (OR_ZEBRA_=0.91, p=0.0099 / OR_MCV_=1.11, p=0.0054). GRS_EBNA_ was associated with both increased risk for *IDH* mutated gliomas (OR=1.09, p=0.040) and improved survival (HR=0.86, p=0.010). *HLA-DQA1*03:01* was significantly associated with decreased risk of glioma overall (OR=0.85, p=3.96×10^−4^) after multiple testing adjustment.

This first systematic investigation of the role of genetic determinants of viral antigen reactivity in glioma risk and survival provides insight into complex immunogenomic mechanisms of glioma pathogenesis. These results may inform applications of antiviral based therapies in glioma treatment.

## Introduction

Studies linking viruses and cancer date back over 100 years^1^ and laid the foundation for understanding oncogenes^2^. It has also become increasingly clear, as well evidenced by the current pandemic, that host genetics play an important role in response to viruses^3,4^. To date, seven viruses have been accepted to be tumor-initiating in humans: Epstein-Barr virus (EBV), hepatitis B virus (HBV), human papillomavirus (HPV), human T-lymphotropic virus-1 (HTLV-1), hepatitis C (HCV), Kaposi’s sarcoma herpesvirus (HHV-8) and Merkel cell polyomavirus (MCV)^5^. Recent analyses have shown that infections account for approximately 13% of human cancers worldwide^6^. However viruses have not been definitively implicated in the etiology of glioma despite decades of suggestive associations^7–11^.

Glioma is a highly fatal brain cancer with a paucity of known risk factors, with exposure to ionizing radiation being the accepted causal factor^12^. A history of previous infection with Varicella-Zoster virus (VZV) has been the only infectious agent consistently linked to adult glioma, conferring an estimated 20% decrease in risk^10,13^. A suite of other viruses have been associated with the risk and grade of glioma, including EBV, MCV, John Cunningham virus (JCV), BK virus (BKV), human Cytomegalovirus (CMV) and human herpesvirus-6 (HHV-6), but with discordant results^11,14–16^. The identification of key somatic molecular alterations (e.g. *IDH* mutation, 1p/19q chromosomal arm codeletion, *TERT* mutation), which drastically affect glioma prognosis, have uncovered subtype-specific risk factors^17,18^. Recently, several studies/clinical trials have suggested a prognostic benefit of antiviral medications in the treatment of glioma^19–21^.

Genetic host response to viral infection could play a role in elucidating potential links between viruses and cancer incidence and prognosis. Studies have demonstrated significant heritable components (32-48%) of antibody response to many viruses and have identified genetic loci within host genes related to cell entry, cytokine production, and immune response^22–24^. Genetic variants of class I and II human leukocyte antigen (HLA) genes contribute the most important identified components of genetic determinants of response to viral antigens. Class II genes each encode half of a heterodimeric class II HLA protein, which presents extra-cellularly-derived peptides to CD4+ helper T-cells. The immune response is triggered when a CD4+ T-cell recognizes the combination of a call II HLA protein and its bound peptide. It is well studied that CD4+ T-cells have an important role in creating and sustaining effective anti-tumor immunity^25^.

The HLA region of the genome is considered the most polymorphic region of the human genetic system^26^ where polymorphisms have been shown to alter the risk and progression of disease in a variety of autoimmune (notably HLA class II) and malignant conditions^27,28^. Certain HLA haplotypes have shown non-additive epistatic effects on glioma risk^29^, yet no germline variants within the HLA have been directly identified as risk loci in a glioma GWAS.

In this study we leveraged previously published genome-wide SNP associations with viral antibody response^30^ to generate genetically inferred antigen reactivity profiles in glioma cases and controls and evaluated their association with risk and survival by major glioma subtypes. We further conducted imputed HLA gene association analysis with glioma risk and survival and our findings suggest a convergence of genetic mechanisms regulating host immune response to viral challenge and glioma development and progression.

## Methods

### Ethics

Collection of patient samples and associated clinicopathological information was undertaken with written informed consent and relevant ethical review board approval at the respective study centers in accordance with the tenets of the Declaration of Helsinki. Specifically informed consent and ethical board approval was obtained from the UCSF Committee on Human Research (USA) and the Mayo Clinic Office for Human Research Protection (USA). The diagnosis of glioma (ICDO-3 codes 9380-9480 or equivalent) was established through histology in all cases in accordance with World Health Organization guidelines.

### Study Populations

We analyzed three glioma case-control sets assembled based on genotyping platform and study population for a total sample size of 3418 cases and 8156 controls (**Figure 1, Table 1**). The first set included 1973 cases from the Mayo Clinic and University of California, San Francisco (UCSF) Adult Glioma Study and 1859 controls from the Glioma International Case-Control Study (GICC) who were genotyped on the Illumina OncoArray, as previously described^18,31–34^. The second dataset included 659 cases and 586 controls from the UCSF Adult Glioma study genotyped on the Illumina HumanHap370duo panel^32^. The third dataset included 786 glioma cases from The Cancer Genome Atlas (TCGA) with available molecular data genotyped on the Affymetrix 6.0 array. Cancer-free controls were assembled from two Wellcome Trust Case Control Consortium (WTCCC) studies genotyped using the Affymetrix 6.0 array: 2,917 controls from the 1958 British Birth cohort and 2,794 controls from the UK Blood Service control group.

**Figure 1:**
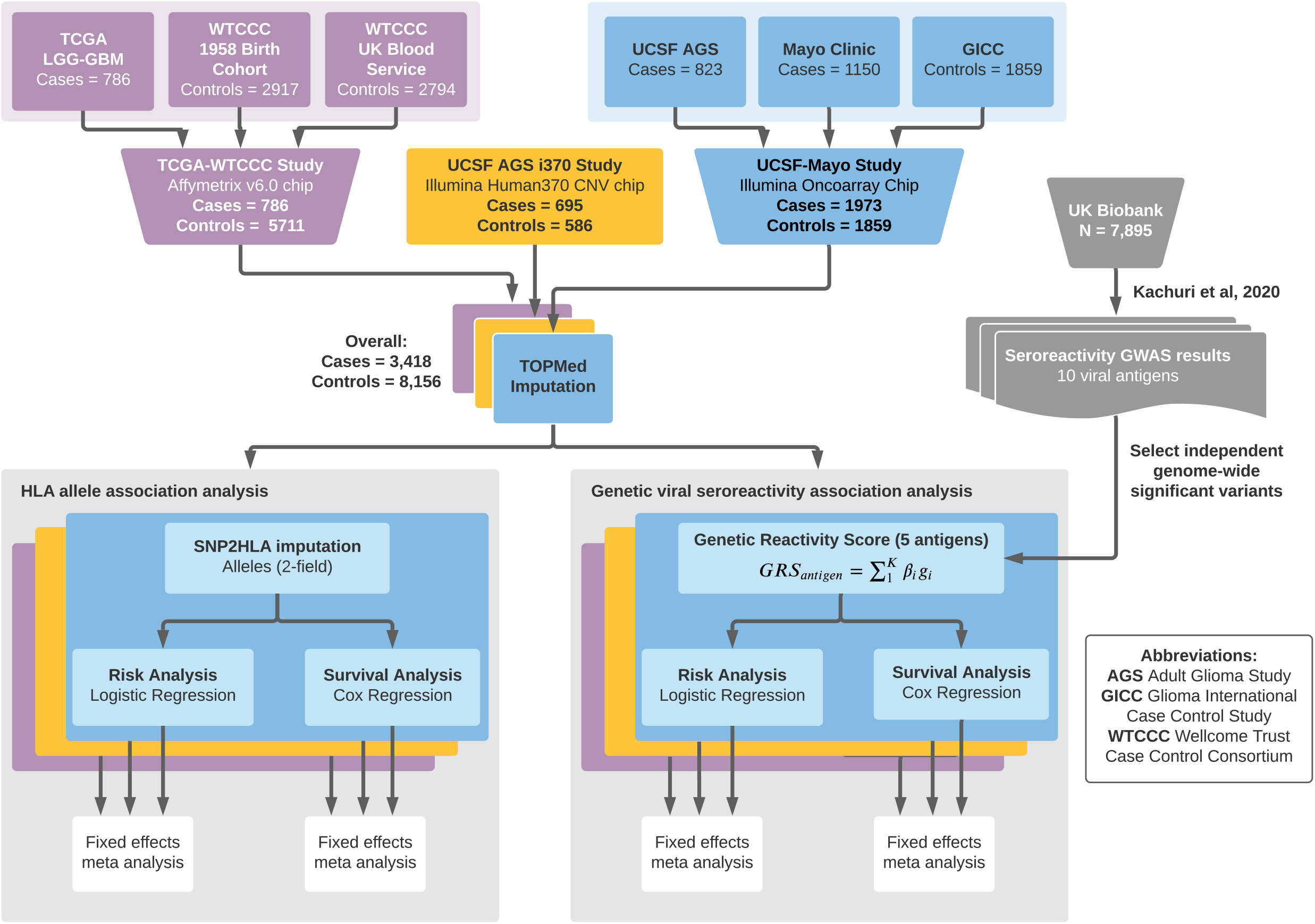
Summary of data processing and analysis. Our analysis consisted of three glioma case-control datasets. The first dataset (purple) included 786 glioma cases from The Cancer Genome Atlas (TCGA) genotyped on the Affymetrix 6.0 array and cancer-free controls assembled from two Wellcome Trust Case Control Consortium (WTCCC) studies genotyped using the Affymetrix 6.0 array: 2,917 controls from the 1958 British Birth cohort and 2,794 controls from the UK Blood Service control group. The second dataset (yellow) included 659 cases and 586 controls from the University of California, San Francisco (UCSF) Adult Glioma Study (AGS) genotyped on the Illumina HumanHap370duo panel^32^. The third set (blue) included 1973 cases from the Mayo Clinic and UCSF AGS and 1859 controls from the Glioma International Case-Control Study (GICC) who were genotyped on the Illumina OncoArray, as previously described^18,31–34^. The three resulting case-control datasets were processed through quality controls as described in the main text and imputed using the TOPMed imputation server. SNP2HLA was used to impute HLA alleles from SNP data. Risk and survival analysis were performed separately on each study’s imputed HLA alleles, on multiple glioma molecular subtypes, and a fixed-effects meta analysis was performed to aggregate results. Separately, genetic reactivity scores to 5 viral antigens were created using previously published GWAS data. Cases and controls across the 3 studies were then assessed a genetically predicted antibody reactivity score (GRS) to each of the 5 antigens. Risk and survival analysis were performed separately on each study using each GRS score as a predictor, with results aggregated using a fixed effects inverse variance weighted meta analysis for each subset of glioma patients and controls based on molecular subtype.

**Table 1:** Clinical and molecular summary of the three case-control datasets.

|  | UCSF-Mayo<br>Cases<br>N=1973 | GICC<br>Controls<br>N=1859 | AGS i370<br>Cases<br>N=659 | AGS i370<br>Controls<br>N=586 | TCGA<br>Cases<br>N=786 | WTCCC<br>Controls<br>N=5711 |
| --- | --- | --- | --- | --- | --- | --- |
| Age |  |  |  |  |  |  |
| <40 | 983 (50%) | 538 (29%) | 101 (15%) | 85 (15%) | 213 (27%) | 0 (0%) |
| 40-59 | 476 (24%) | 555 (30%) | 329 (50%) | 252 (43%) | 279 (36%) | 0 (0%) |
| >= 60 | 514 (26%) | 766 (41%) | 229 (35%) | 249 (42%) | 245 (31%) | 0 (0%) |
| Missing | 0 (0%) | 0 (0%) | 0 (0%) | 0 (0%) | 49 (6%) | 5711 (100%) |
| Sex |  |  |  |  |  |  |
| Female | 822 (42%) | 744 (40%) | 229 (35%) | 280 (48%) | 328 (42%) | 2819 (49%) |
| Male | 1151 (58%) | 1115 (60%) | 430 (65%) | 306 (52%) | 458 (58%) | 2892 (51%) |
| IDH mutation status |  |  |  |  |  |  |
| Mutant | 588 (30%) | NA | 111 (17%) | NA | 375 (48%) | NA |
| Wild type | 699 (35%) | NA | 416 (63%) | NA | 364 (46%) | NA |
| Missing | 686 (35%) | NA | 132 (20%) | NA | 47 (6%) | NA |
| Molecular subtype based on IDH mutation and 1p/19q codeletion |  |  |  |  |  |  |
| MT-codel | 244 (12%) | NA | 9 (1%) | NA | 143 (18%) | NA |
| MT-noncodel | 291 (15%) | NA | 94 (14%) | NA | 230 (29%) | NA |
| WT-noncodel | 507 (26%) | NA | 416 (63%) | NA | 357 (46%) | NA |
| Missing/other | 905 (47%) | NA | 140 (22%) | NA | 56 (7%) | NA |

NA: not applicable to controls

Molecular subtype information (*IDH* mutation and 1p/19q codeletion status) was downloaded from Ceccarelli et al.^35^ supplementary table 1 for the third dataset and was provided directly from the UCSF AGS study and Mayo Clinic for the first and second datasets.

### Quality Control and Imputation

Standard quality control procedures were implemented prior to imputation. Analyses were restricted to individuals of predominantly (>70%) European ancestry, determined using ADMIXTURE^36^ and the HapMap 3 reference populations. Within each ancestral group, we removed samples with excess heterozygosity (>3 standard deviations (SD) from mean), <95% call rates, and discordant self-reported and genetically inferred sex. Relatedness checks were performed within each study using KING^37^ (kinship >0.12), filtering out up to second-degree relations and retaining the samples with higher call rate. TCGA blood samples were preferentially chosen when both blood and tumor sequencing data were available for the same patient (693 European samples had both blood and tumor samples available). SNPs with <95% call rate were removed, along with variants deviating from Hardy-Weinberg equilibrium (p<10^−6^) or at a low minor allele frequency (MAF<0.005). Samples genotyped on the same platform (i.e., Affymetrix 6.0 for TCGA and WTCCC) were imputed together using the multi-ethnic TOPMed reference panel (ver. r2).

## Statistical Analysis

### Genetically Predicted Viral Antigen Response

Genetic seroreactivity scores (GRS) were calculated to obtain genetically inferred viral antigen response profiles in each of the glioma datasets. For each GRS, candidate variants and corresponding effect sizes were obtained from genome-wide summary statistics for seroreactivity to 10 viral antigens previously identified in Kachuri and Francis et al. 2020^30^ in 7895 randomly selected individuals of European descent from the UK Biobank (UKB) cohort using serological measures. For each antigen, we preferentially selected independent SNPs (linkage disequilibrium, LD, r^2^<0.01 within 500kb) with the lowest p-value among genome-wide significant variants (p<5×10^−8^). LD proxies (r^2^>0.9) were obtained for variants unavailable in the target glioma datasets. For each individual, an antigen-specific GRS was calculated as a weighted sum with weights (β) corresponding to a standard deviation increase in antibody response:

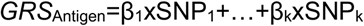

Each GRS was calculated using a minimum of 4 variants with MAF>=0.01 and imputation quality (R^2^>0.6), and then standardized within each dataset.

#### Antigen GRS Associations with Glioma Susceptibility

GRS associations with disease risk were examined for glioma overall and for molecular subtypes defined by the specific tumor alterations: *IDH* mutation status and 1p/19q chromosomal arm codeletion. For each GRS, odds ratios (OR) were estimated using logistic regression models adjusted for age, sex, and the first 10 genetic ancestry principal components (PCs). Models in the UCSF-Mayo dataset were further adjusted for contributing site. GRS associations from each study were meta-analyzed using a fixed-effects inverse-variance weighted approach. Heterogeneity in study-specific GRS associations was assessed using Cochran’s Q test.

#### Antigen GRS Associations with Glioma Survival

The association between each antigen-GRS and overall survival was assessed using a Cox proportional hazards regression model with follow-up time calculated from the date of first surgery to either date of death or last known contact. The latter censored at that date. Analyses were conducted for glioma overall and molecular subtypes with a minimum of 50 cases and 20 events (deaths). Proportionality assumptions were checked within each dataset via examination of Kaplan-Meier curves. Hazard ratios (HR) were estimated using Cox models adjusted for age, sex, 10 genetic PCs, and study site (if applicable). Associations with survival in each study were combined using fixed-effects meta-analysis. For each nominally significant (p<0.05) glioma-GRS association, survival differences were further assessed using Kaplan-Meier curves by comparing mortality trajectories in patients with high genetically predicted immune reactivity (top 20%) to the remainder.

#### Regional HLA Analyses of glioma risk and survival

Classical HLA alleles were imputed for samples in all cohorts at two-field resolution using SNP2HLA^38^ and the Type 1 Diabetes Genetics Consortium (T1DGC) reference panel of 2,767 unrelated individuals. Associations were tested for 77 alleles (MAF>=0.01) with imputation quality >0.4 across eight genes: *HLA-A, HLA-B, HLA-C, HLA-DPA1, HLA-DPB1, HLA-DQA1, HLA-DQB1, HLA-DRB1*.

Subtype-specific associations with risk were estimated using logistic regression models. HLA allele associations with mortality were assessed using SPACox^39^, an extension of the Cox model with improved type I error control in high-dimensional settings. Study-specific risk and survival associations were combined in a meta-analysis. Associations for each HLA allele were considered statistically significant if p<6.5×10^−4^ (0.05/77 alleles).

## Results

The creation of a GRS was attempted for all antigens with genome-wide significant seroreactivity-associated variants (p<5×10^−8^) as reported in Kachuri and Francis et al. 2020^30^. This included 10 antigens: Four EBV antigens (EA-D, EBNA, p18, ZEBRA) and antigens for BKV, HHV7, HSV1, JCV, MCV, and VZV. We do not include antigens for CMV and HHV6 in our study as there was only one significantly associated SNP each in the previously published GWAS. An antigen-specific GRS was considered successfully created if at least four independent SNPs passed LD clumping and thresholding and were present (or via proxy SNP) in the three glioma study datasets. GRS were successfully generated for EBV EA-D, EBV EBNA, EBV p18, EBV ZEBRA, and MCV. Only one independent SNP remained for each of HSV1, BKV, JCV and VZV after accounting for long-range LD in the HLA region, although having numerous SNPs meet genome-wide significance in the previous seroreactivity study. Three SNPS remained for HHV7, each on a different chromosome (6,11, and 17). Complete information of variants considered for all 12 viral antigens, including nearest gene and corresponding glioma risk associations are reported in **Supplement Tables S1 and S2**. LD correlations for all associated chromosome 6 variants are available in a heatmap in **Supplement Figure S1**.

The variants included in each of the five created GRSs were overwhelmingly located in the HLA region, with few predictors located elsewhere across the genome: rs67886110 in 3q25.1 an eQTL for *MED12L* and *P2RY12* (EBV EBNA), rs7618405 in 3p24.3 (MCV), and rs7444313 in 5q31.2 near *TMEM173* (MCV). Of the 38 SNPs included across the 5 GRSs, 2 had a significant association (Bonferroni-corrected: p<1.3×10^−3^) with overall glioma risk: rs9265517 near *HLA-B* (EBV EBNA), p=3.08×10^−4^ and rs9268847 near *HLA-DRB9* (MCV), p=2.86×10^−4^. **Supplement Table S2** visualizes the squared correlation between the GRS SNPs and each imputed HLA allele (two field resolution) from UCSF-Mayo cases and controls. Notably, the GRS for MCV was inversely correlated with GRSs for EBV ZEBRA (Pearson’s r=-0.28, p=3.89×10^−70^) and EBV EBNA (r=-0.19, p=6.28×10^−31^) (**Figure 2)**.

**Figure 2:**
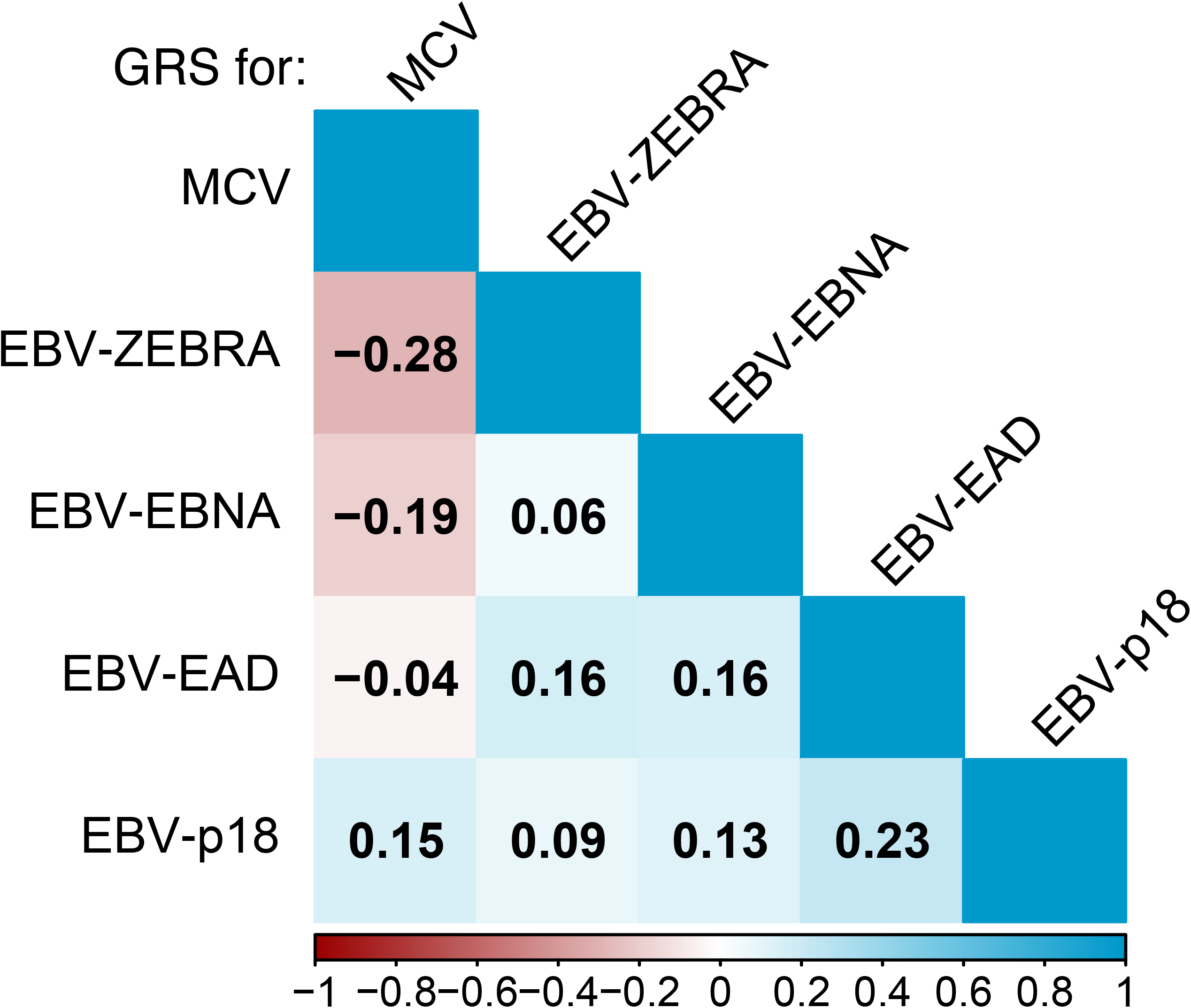
GRS correlations within the UCSF-Mayo cases and controls. Pearson correlations between genetically predicted antigen responses (via GRS) as computed in the UCSF-Mayo glioma cases and controls. Values were printed in each block if and only if the associated correlation test p-value was less than 0.01.

### Viral antigen GRS associations with glioma risk

In the combined meta-analysis of three case-control studies, three GRS’s reached at least nominal significance (p<0.05), with some associations remaining statistically significant after correction for the number of antigens tested (Bonferroni: p=0.05/5=0.01) (**Figure 3**). Genetic predisposition to an increased serological response to EBV ZEBRA was inversely associated with risk of glioma overall (per 1 SD increase in GRS: Odds Ratio, OR_ZEBRA_=0.94, 95% confidence interval 0.89-0.99, p=0.012, 3418 cases). GRS_EBNA_ was associated with an increased risk of *IDH* mutated gliomas (OR_EBNA_=1.09,1.004-1.18, p=0.040, 1074 cases) and the magnitude of this effect increased for *IDH* mutated 1p/19q codeleted gliomas (OR_EBNA_=1.14, 1.012-1.28, p=0.031, 396 cases).

**Figure 3:**
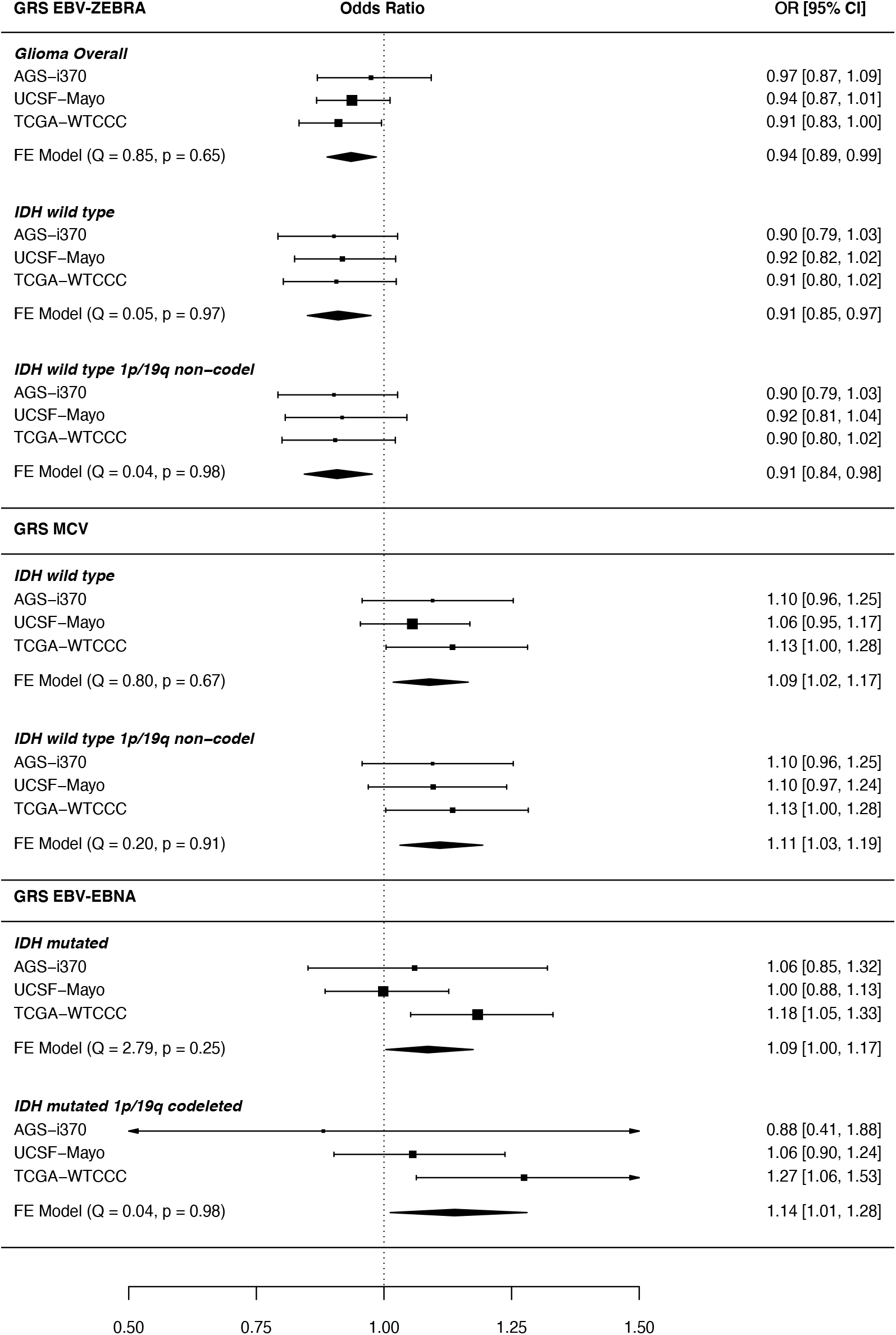
Significant GRS-glioma subtype risk association meta-analysis forest plots. Forest plot meta-analysis results of GRS-glioma risk associations which were at least nominally statistically significant (p <0.05). Response to antigens EBV ZEBRA, MCV and EBV EBNA had associations which reached this threshold. Results are reported as odds ratios, along with 95% confidence intervals. Briefly, each header indicates the studied viral antigen GRS, within are its association with molecular glioma subtypes reported with p<0.05, and the 95% confidence interval of each study-specific effect. The diamond visualizes the 95% confidence interval for the fixed effect (FE) meta-analysis across all 3 studies. Each meta-analysis was tested for between-study heterogeneity (Q statistic), with p<0.05 indicating evidence of study-specific associations.

We observed some evidence of antagonistic pleiotropy between genetic determinants of antibody response to EBV ZEBRA and MCV, which generalized across glioma subtypes. GRS_ZEBRA_ and GRS_MCV_ were associated with susceptibility to *IDH* wild type gliomas, but in opposite directions: Higher genetically predicted reactivity to EBV ZEBRA was inversely associated with *IDH* wild type glioma risk (OR_ZEBRA_=0.91, 0.85-0.98, p=0.0072, 1479 cases), while increased predicted antibody response to MCV conferred an increased risk (OR_MCV_=1.09, 1.02-1.17, p=0.013). This pattern persisted for *IDH* wild type 1p/19q non-codeleted gliomas (OR_ZEBRA_=0.91, 0.84-0.9, p=0.0099; OR_MCV_=1.11, 1.03-1.19, p=0.0054, 1280 cases). The correlation between GRS_ZEBRA_ and GRS_MCV_ (Pearson’s r=-0.28, p=3.9×10^−70^ in UCSF-Mayo, **Figure 2**) suggests a possible shared underlying genetic mechanism between the response to the two antigens. GRS_ZEBRA_ and GRS_MCV_ were tested together, along with interaction term, in a single logistic model and we observed no significant interaction (p=0.34 in *IDH* wild type risk).

### Viral antigen GRS-survival associations

The number of available cases and deaths across the three studies is available in **Table 2**. Associations between genetically predicted viral antigen response profiles and survival were restricted to *IDH* mutated gliomas (**Figure 4**). GRS_EBNA_ was associated with survival in 1074 *IDH* mutated glioma cases (per 1 SD increase: Hazard Ratio, HR=0.86, 0.76-0.96, p=0.010, 325 events), suggesting that a higher genetically predicted reactivity to EBV EBNA improved duration of survival.

**Table 2:**
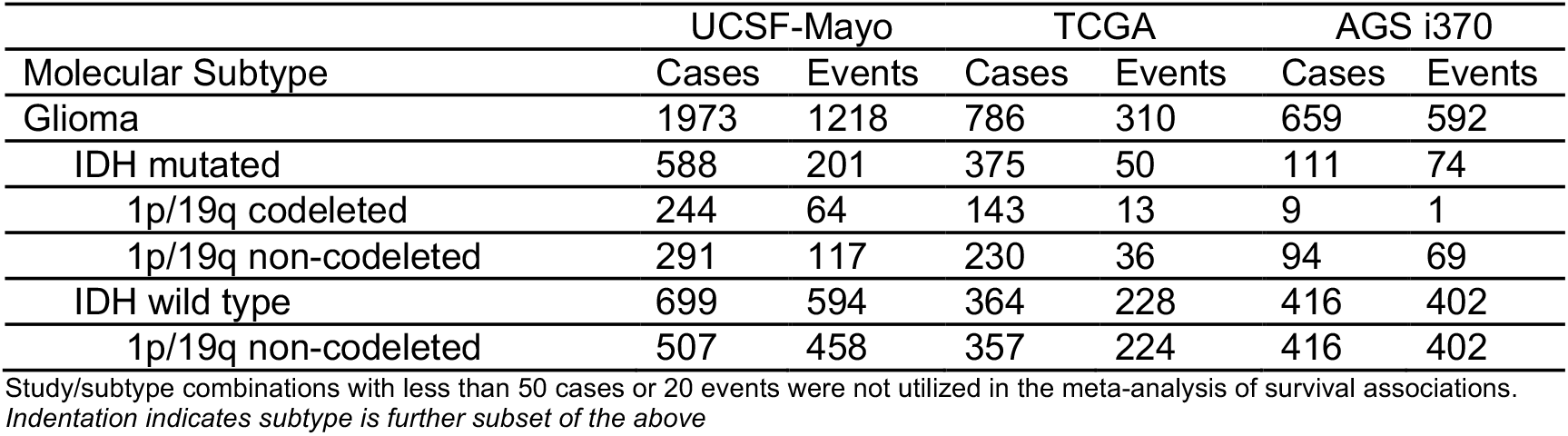
Summary of available cases and events for survival analyses.

|  | UCSF-Mayo |  | TCGA |  | AGS i370 |  |
| --- | --- | --- | --- | --- | --- | --- |
| Molecular Subtype | Cases | Events | Cases | Events | Cases | Events |
| Glioma | 1973 | 1218 | 786 | 310 | 659 | 592 |
| IDH mutated | 588 | 201 | 375 | 50 | 111 | 74 |
| 1p/19q codeleted | 244 | 64 | 143 | 13 | 9 | 1 |
| 1p/19q non-codeleted | 291 | 117 | 230 | 36 | 94 | 69 |
| IDH wild type | 699 | 594 | 364 | 228 | 416 | 402 |
| 1p/19q non-codeleted | 507 | 458 | 357 | 224 | 416 | 402 |

Study/subtype combinations with less than 50 cases or 20 events were not utilized in the meta-analysis of survival associations. *Indentation indicates subtype is further subset of the above*

**Figure 4:**
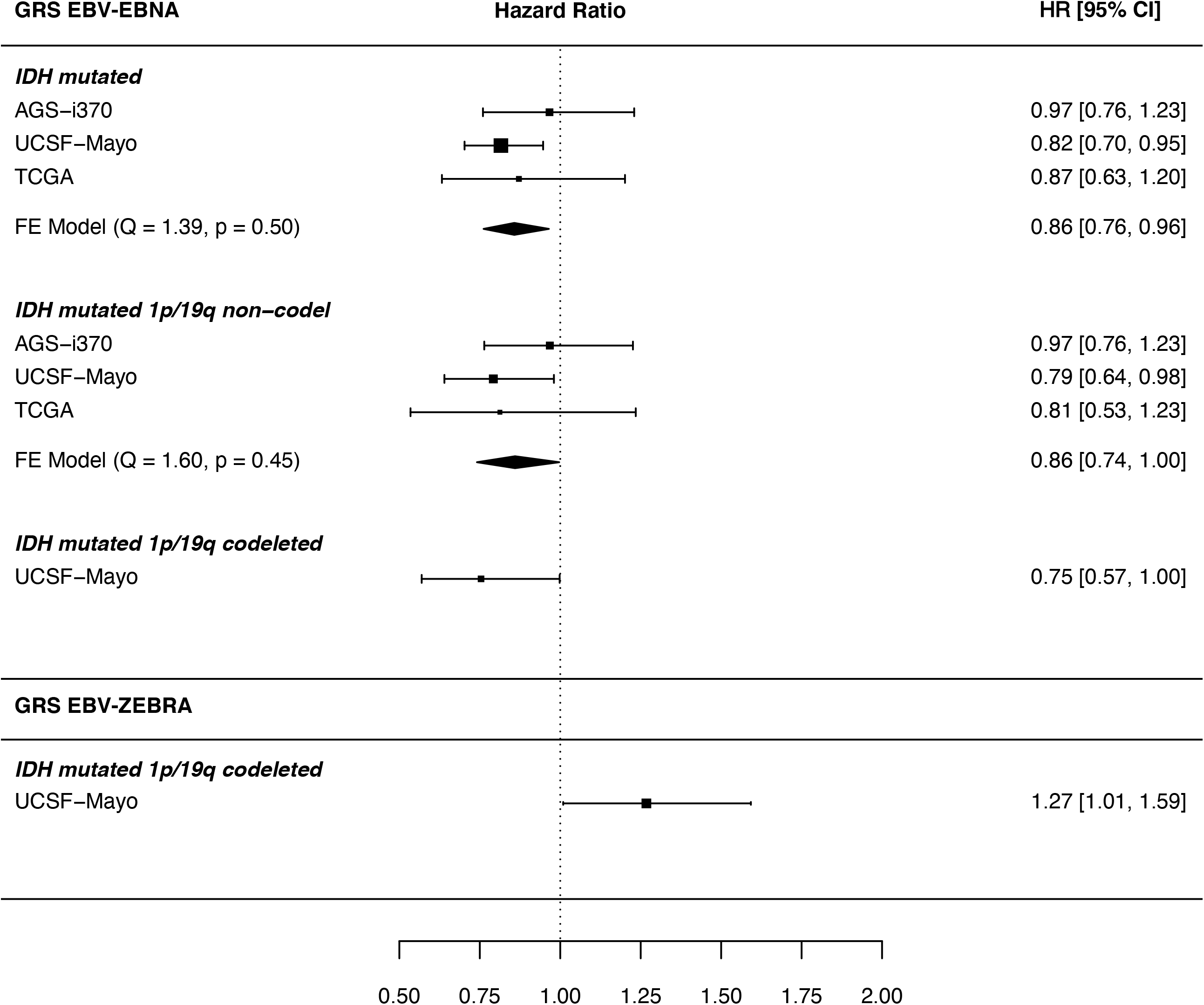
Nominal GRS-glioma subtype survival association meta-analysis forest plots. Forest plot meta-analysis results of GRS-glioma survival associations which were at least nominally statistically significant (p <0.05). Genetically inferred response to antigens EBV ZEBRA, MCV and EBV EBNA had associations which reached this threshold. Results are reported as hazard ratios, along with 95% confidence intervals. Briefly, each header indicates the studied viral antigen GRS, within are its association with molecular glioma subtypes reported with p<0.05, and the 95% confidence interval of each study-specific effect. The diamond visualizes the 95% confidence interval for the fixed effect (FE) meta-analysis across included studies. Studies which had an insufficient number of cases/events in a subtype were not included in the meta-analysis. Each meta-analysis (where more than one study was included) was tested for between-study heterogeneity (Q statistic), with p<0.05 indicating evidence of study-specific associations.

GRS for two EBV antigens were nominally associated with survival amongst 244 *IDH* mutated 1p/19q codeleted glioma cases, but in opposite directions (HR_EBNA_=0.75, 0.57-0.99 p=0.048 / HR_ZEBRA_=1.27,1.01-1.6, p=0.042, 64 events). This subtype-specific result was limited to the UCSF-Mayo dataset due to insufficient number of reported events (deaths) in *IDH* mutated 1p/19q codeleted glioma cases from TCGA and AGS-i370.

GRS_EBNA_ was also associated with improved survival outcomes in 614 *IDH* mutated 1p/19q non-codeleted glioma cases (HR=0.75, 0.74-0.997, p=0.045, 222 events), suggesting the association of GRS_EBNA_ is independent of 1p/19q status. **Figure 5** visualizes the significant GRS_EBNA_ associations using Kaplan-Meier survival curves.

**Figure 5:**
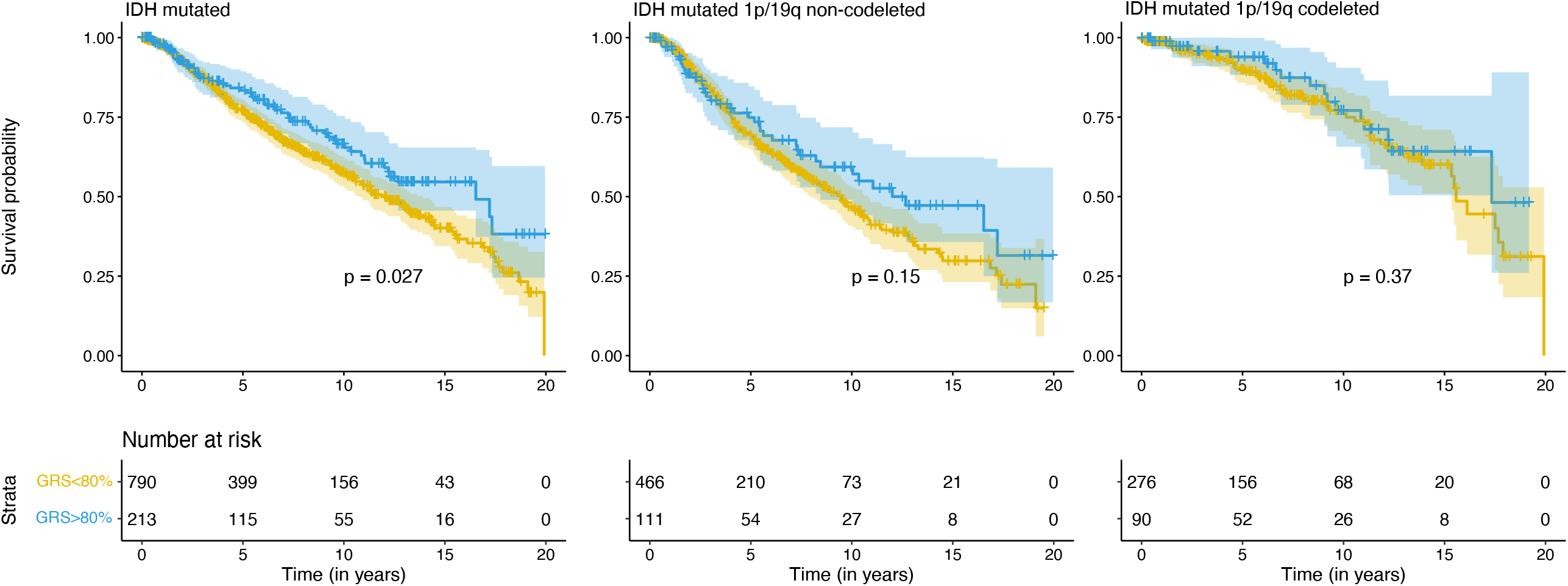
Kaplan Meier curves for significant GRS_EBNA_ - glioma molecular subtype associations. Kaplan-Meier curves for subtypes where GRS_EBNA_ was nominally associated with subtype specific glioma survival differences (p <0.05 via Cox regression). The second and third plots are distinct partitions of the IDH mutated subgroup. To visualize, unnormalized GRS scores across the included studies were binned based on the case-specific 80^th^ percentile score in the UCSF-Mayo dataset. P-values included on each plot are results of a log-rank test for difference between the two curves. Below each set of curves provides the number of cases surviving beyond that time point, for each of the two GRS groups. In all cases, the glioma cases with higher GRS score for EBV EBNA had visually improved survival outcomes compared to the bottom 80%.

We did not detect any significant GRS associations in the analysis of glioma overall, which is consistent with the strong prognostic significance of molecular glioma subtypes^17,33^. Also, as the prognosis in *IDH* wild type gliomas is the poorest, we suspect our GRS instruments are underpowered to detect significant deviations over such short intervals. Full survival results are available in **Supplement Figures S3 and S4**.

### HLA allele-glioma associations

Of the 77 HLA alleles imputed at two-field resolution, only *HLA-DQA1*03:01* reached Bonferroni-corrected significance (p<6.5×10^−4^ after correcting for 77 HLA alleles tested) for association with glioma risk in a meta-analysis. The presence of an *HLA-DQA1*03:01* allele was associated with a decrease in overall glioma risk (OR=0.85, p=3.96×10^−4^). This allele was also nominally associated with risk of *IDH* wild type (OR=0.82, p=1.4×10^−3^) and *IDH* wild type 1p/19q non-codeleted gliomas (OR=0.81, p=2.6×10^−3^). These subtype-specific associations and direction of effect mirror those of GRS_ZEBRA_ presented above, suggesting *HLA-DQA1*03:01* as a potential shared marker for both glioma risk and EBV ZEBRA seroreactivity. Results of all HLA allele (one and two field resolution) associations with glioma risk tested in a meta-analysis are available in **Supplement Table S2**.

## Discussion

We investigated genetically predicted antibody response to ten antigens for seven viruses in relation to glioma risk and prognosis in a meta-analysis of three patient cohorts with molecular subtype information. Our analysis discovered evidence that genetic predisposition to reactivity to specific viral antigens associated with susceptibility to glioma in a subtype-specific manner. This pattern of effect modification by glioma subtype also extends into prognosis, where we observed associations between viral antigen GRSs and survival among specific glioma subtypes.

One of our main findings is that genetic predisposition to increased seroreactivity to EBV ZEBRA was associated with a decreased overall glioma risk, with a significant decrease in *IDH* wild type subtypes. Predicted reactivity to the MCV VP1 antigen mirrored the same *IDH* wild type associations but in the opposite direction, where higher reactivity was associated with increased glioma risk. The significant inverse relationship between predicted reactivity to EBV ZEBRA and MCV VP1 highlights the possibility of shared genetic components of antibody response to the two antigens. Understanding the underlying mechanisms guided by these genetics is left as an open question. A possible link is that the class II HLA allele *DQA1*03:01* was associated with decreased glioma risk in the same subtypes associated with GRSs for EBV ZEBRA. In our previous UKB analysis^30^ the presence of *HLA-DQA1*03:01* was positively associated with EBV ZEBRA seroreactivity measurements (β=0.168, p=1.3×10^−16^). Taken together, the associations between *HLA-DQA1*03:01*, EBV ZEBRA and glioma risk suggest possible shared underlying immunogenetic architecture. As HLA class II proteins present potentially antigenic peptides, functional genetic alterations can result in changes to the binding affinity of specific antigens, modulating the potential for recognition by CD4+ helper T-cells. It is possible that the heterodimeric DQ proteins half-encoded by *DQA1*03:01* have improved binding or recognition of peptides presented by both EBV ZEBRA proteins and glioma (specifically *IDH* wild type), allowing for an increased immune response in both cases. As *IDH* wild type has generally been shown to serve as a marker for more severe glioma cases, the exact somatic tumor alterations leading to recognized peptide variants in these tumors is not clear and is an area that warrants future investigation. Further analysis of viral-tumor protein homology is needed to understand if this could be a possible connection.

Interestingly, a higher genetically predicted response to EBV EBNA was nominally associated with increased risk in *IDH* mutated/1p/19q codeleted gliomas. Still, reactivity towards EBV EBNA was more strongly associated with improved survival in *IDH* mutated gliomas. This discrepancy between the disease-promoting and pro-survival associations of GRS_EBNA_ may suggest the presence of different biologic pathways after initiation of disease. It may also point towards EBV latency-mediated treatment effects. Previous studies show that the EBV latency type predicts the relative amount of induced reactivation generated by cytotoxic chemotherapy drugs^40^. Therefore, individuals who react strongly to EBV EBNA antigens may exhibit a different pattern of reactivation when treated with temozolomide. Further research is required to elucidate this putative association, yet it is clear that EBV lytic/latent cycling is a critical aspect of other germline interactions and disease risk.

Epstein-Barr virus was the first recognized human oncovirus^41^ and exists in two distinct life cycles within a host: a lytic phase of active infection where new viruses are produced, and a latent phase where the virus remains dormant to avoid detection by the host. Distinct sets of antigens are produced during these two phases, two of such are EBNA, during the latent phase of infection, and the ZEBRA protein which initiates a change from latent to lytic gene expression. Past evidence suggests processes during the latent stage are responsible for the virus’ oncogenic properties via mechanisms promoting cell growth and preventing cell apoptosis (reviewed in Akhtar et al. 2018^42^). Recent work has also implicated the ZEBRA protein as oncogenic with evidence demonstrating its ability to deregulate immune surveillance and promote immune escape^43^.

In contrast MCV is the most recently recognized human oncovirus, first described in 2008 and later accepted as a causal agent for Merkel cell carcinoma^44^, a neuroendocrine carcinoma. Previous studies have identified the presence of MCV DNA in glioma patients and have drawn an association with infection and increased risk of glioma^15,16^. It has been shown that both the large T and small T antigens of MCV are oncoproteins that target tumor suppressor proteins such as pRB. Although the exact latency mechanism in polyomaviruses is unknown, it has been suggested that complex protein-mediated latency may be critical to the MCV lifecycle^45^.

In interpreting our findings, several limitations should be acknowledged. The shared genetic architecture between the viruses studied here, as seen in the correlations in **Figure 2 and Supplement Figure S1**, may result in a lack of specificity. This limitation in our study may be applicable in other viruses that share the same underlying genetic programming of antigen response, particularly VZV, which is associated with a unique pattern of LD spanning a large region of the HLA^30^. Furthermore, the clumping and thresholding approach to GRS development may not be optimal for regions with complex LD structure, such as HLA. An approach that can appropriately account for the long-range correlation structure and capture non-linear interactions, such as haplotype effects, may improve future studies that use genetically inferred immune responses. Another challenge is the low SNP density in the HLA region in the present study, as well as reliance on imputed HLA alleles both in the glioma studies and in UKB. The HLA region is highly polymorphic and imputation of alleles from SNP data is limiting and prone to error. Similarly, it is difficult to disentangle the specific function of SNPs located in the HLA region. As such, additional studies leveraging HLA sequencing approaches are required to provide further granularity of the genetic determinants of antigen response and glioma risk/survival.

Additionally, we could not study the relationship between reactivity to several antigens of interest (BKV, HSV-1, JCV, VZV, HHV-7, HHV-6, CMV) due to the limited number of variants reaching genome-wide significance and/or meeting our GRS criteria^30^. Although the GRS-glioma association results presented here require further replication in independent patient populations, our findings are intriguing and suggest previous inconsistencies in observational epidemiologic may be partially due to individual differences in genetic factors that affect antigen reactivity.

This is the first study directly examining the underlying genetic architecture of antigen reactivity to common viruses and glioma risk and survival. We observed important associations between programmed reactivity to viruses and glioma etiology and prognosis. This methodology is not limited to the study of glioma, as the GRS instruments proposed here can readily be applied to cohorts of any cancer type. This offers a unique approach for future studies to reinvestigate the genetic contributions of long-running epidemiologic associations between viruses and cancer, and possibly clarify effects of viral therapies in their treatment.

## Data Availability

Genotype data of control samples from the 1958 British Birth Cohort and UK Blood Service Control Group were made available from the Wellcome Trust Case Control Consortium (WTCCC) and downloaded from the European Genotype Archive under ascension numbers EGAD00000000021 and EGAD00000000023, respectively. Genotype data of glioma cases from The Cancer Genome Atlas (TCGA) were obtained from Database of Genotypes and Phenotypes (dbGaP) (phs000178). Genotype data of control samples from the Glioma International Case Control Study (GICC) are available from dbGaP under accession phs001319.v1.p1.

## Supplemental Information

Supplemental data include five tables and five figures.

## Funding and Acknowledgments

Work at University of California, San Francisco was supported by the National Institutes of Health (grant numbers T32CA151022, R01CA52689, P50CA097257, R01CA126831, R01CA139020, R01AI128775, and R25CA112355), the National Brain Tumor Foundation, the Stanley D. Lewis and Virginia S. Lewis Endowed Chair in Brain Tumor Research, the Robert Magnin Newman Endowed Chair in Neuro-oncology, and by donations from families and friends of John Berardi, Helen Glaser, Elvera Olsen, Raymond E. Cooper, and William Martinusen. This publication was supported by the National Center for Research Resources and the National Center for Advancing Translational Sciences, National Institutes of Health, through UCSF-CTSI Grant Number UL1 RR024131. Its contents are solely the responsibility of the authors and do not necessarily represent the official views of the NIH.

The collection of cancer incidence data used in this study was supported by the California Department of Public Health pursuant to California Health and Safety Code Section 103885; Centers for Disease Control and Prevention’s (CDC) National Program of Cancer Registries, under cooperative agreement 5NU58DP006344; the National Cancer Institute’s Surveillance, Epidemiology and End Results Program under contract HHSN261201800032I awarded to the University of California, San Francisco, contract HHSN261201800015I awarded to the University of Southern California, and contract HHSN261201800009I awarded to the Public Health Institute, Cancer Registry of Greater California. The ideas and opinions expressed herein are those of the author(s) and do not necessarily reflect the opinions of the State of California, Department of Public Health, the National Cancer Institute, and the Centers for Disease Control and Prevention or their Contractors and Subcontractors.

## Data Availability

Genotype data of control samples from the 1958 British Birth Cohort and UK Blood Service Control Group were made available from the Wellcome Trust Case Control Consortium (WTCCC) and downloaded from the European Genotype Archive under ascension numbers <u>EGAD00000000021</u> and <u>EGAD00000000023</u>, respectively. Genotype data of glioma cases from The Cancer Genome Atlas (TCGA) were obtained from Database of Genotypes and Phenotypes (dbGaP) (<u>phs000178</u>). Genotype data of control samples from the Glioma International Case Control Study (GICC) are available from dbGaP under accession <u>phs001319.v1.p1</u>.

## Notes

### Authorship

GG, LK, and SSF conceived of the study and wrote main drafts of the manuscript. GG, LK, GW, SJM, NK, and SSF conducted and advised on informatic and statistical analyses along with result interpretations. HMH, AMM, TR, PB, JKW, JEE, RBJ, and MW were involved in primary data collection. All authors contributed and reviewed the final manuscript.

