## Supplemental Tables and Figures for "The immunogenetics of viral antigen response is associated with sub-type specific glioma risk and survival"

#### Table S1: GWAS glioma risk meta results by subtype for the GRS variants

https://ucsf.box.com/s/v2sklvl682f8f8hokzgvh8lzhb7ow9o0

All SNPs that were included in each GRS, basic variant information and beta/SE/p-value for a meta-GWAS for the association of the SNPs for each glioma subtype.

#### Table S2: SNP2HLA HLA variant glioma risk results by subtype

https://ucsf.box.com/s/yusu84xwun539prce7gm7vlowgm027vy

All HLA alleles/genes which were imputed via SNP2HLA, beta/SE/p-value for associations in a meta-analysis of risk analysis of each of the HLA alleles on each glioma subtype.

#### Table S3: Summary of GRS-glioma risk significant results*

| Subtype | Antigen GRS | Meta OR (95% CI) | Meta P-value |
|---|---|---|---|
| Glioma | EBV ZEBRA | 0.936 [0.888, 0.985] | 0.0116 |
| *IDH* mutated | EBV EBNA | 1.086 [1.004, 1.175] | 0.0402 |
| 1p/19q codeleted | EBV EBNA | 1.138 [1.012, 1.280] | 0.0308 |
| *IDH* wild type | EBV ZEBRA | 0.910 [0.850, 0.975] | 0.0072 |
| | MCV | 1.089 [1.018, 1.165] | 0.0131 |
| 1p/19q non-codeleted | EBV ZEBRA | 0.908 [0.844, 0.977] | 0.0099 |
| | MCV | 1.11 [1.031, 1.194] | 0.0054 |

*These results mirror those of Figure 3 in the main text

*Indentation indicates subtype is further subset of the above result*

#### Table S4: Summary of GRS-glioma survival significant results*

| Subtype | Antigen GRS | Meta HR (95% CI) | Meta P-value |
|---|---|---|---|
| *IDH* mutated | EBV EBNA | 0.857 [0.762, 0.964] | 0.010 |
| 1p/19q codeleted | EBV EBNA | 0.754 [0.569, 0.998] | 0.048 |
| | EBV ZEBRA | 1.268 [1.009, 1.592] | 0.042 |
| 1p/19q non-codeleted | EBV EBNA | 0.860 [0.741, 0.997] | 0.045 |

*These results mirror those of Figure 4 in the main text

*Indentation indicates subtype is further subset of the above result*

#### Table S5: Number of SNPs used in each antigen GRS

| | EBV-ZEBRA | EBV-EBNA | EBV-p18 | EBV-EAD | MCV | BKV | HHV7 | HSV1 | JCV | VZV | CMV | HHV6 |
|---|---|---|---|---|---|---|---|---|---|---|---|---|
| **#SNPs** | **11** | **7** | **9** | **5** | **6** | **1** | **3** | **1** | **1** | **1** | **1** | **1** |

Antigens with >4 SNPs were further used in polygenic analyses

#### Figure S1: LD matrix of chromosome 6 SNPs used in GRSs

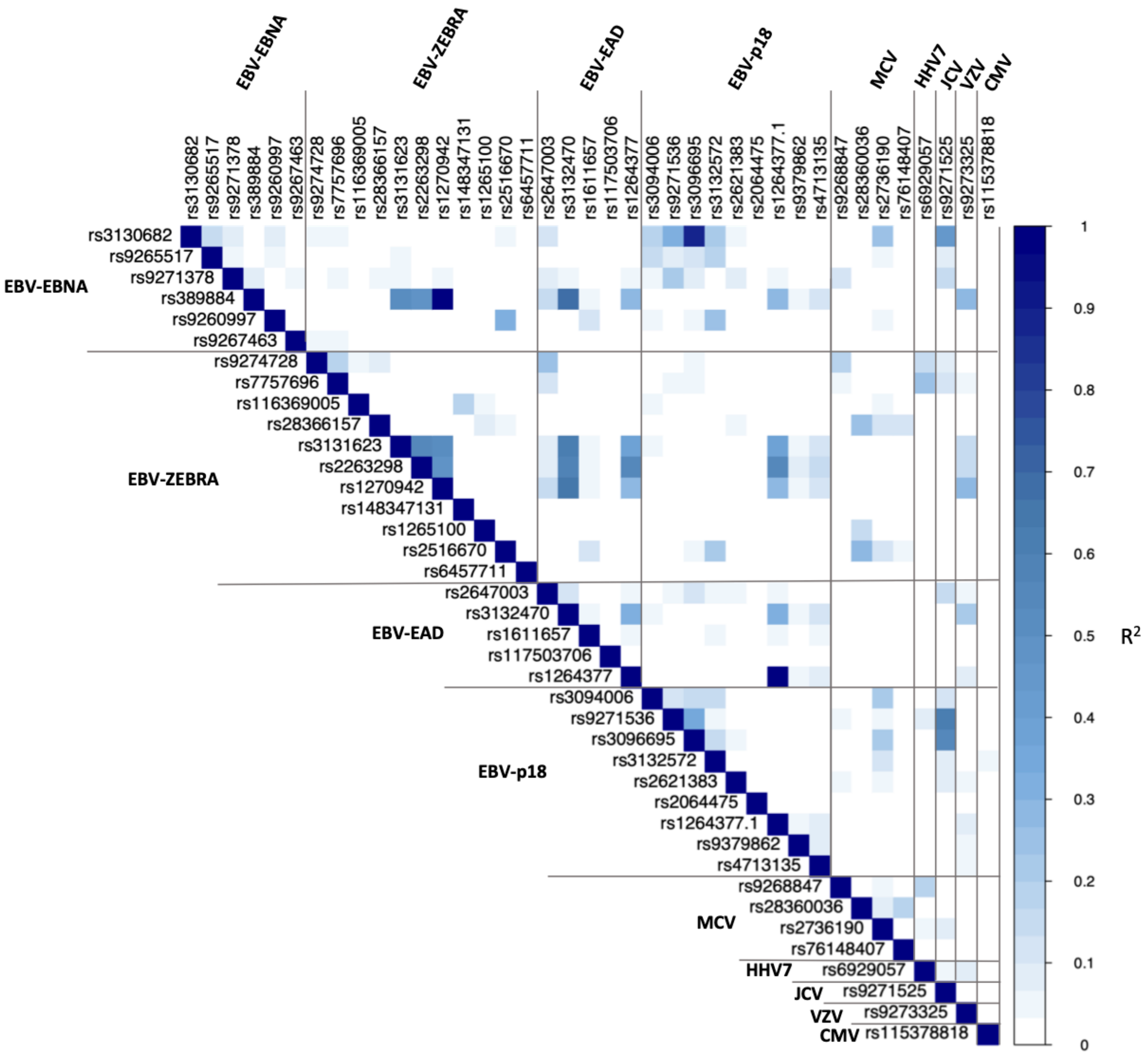


Heatmap of correlation values ($R^2$) of all LD-pruned SNPs on chromosome 6 considered in our GRS analysis, separated by the corresponding significantly associated antigen. Correlations were calculated using LDlink with all available European populations. Cells with darker shades indicate higher levels of correlation/LD.

### Figure S2: $R^2$ matrix of chr 6 GRS SNPs and imputed two-field HLA alleles

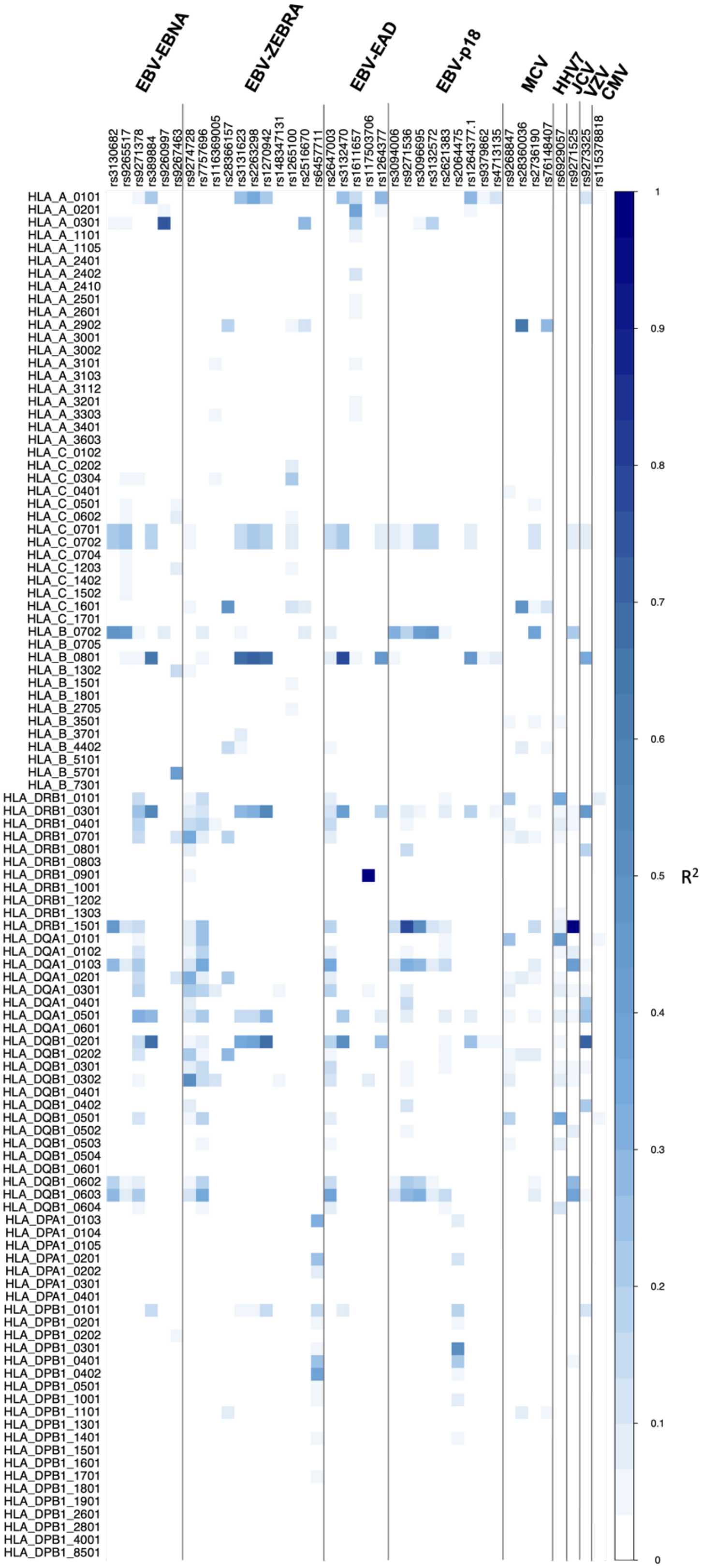

Heatmap of correlation values ($R^2$) of all LD-pruned SNPs on chromosome 6 considered in our GRS analysis, separated by the corresponding significantly associated antigen, with imputed HLA alleles (at two-field resolution). Correlation was calculated using European cases and controls from the UCSF-Mayo dataset. Cells with darker shades indicate higher levels of squared correlation.

### Figure S3: All GRS-Glioma risk meta-result forest plots, by antigen

#### S3 A) Glioma overall

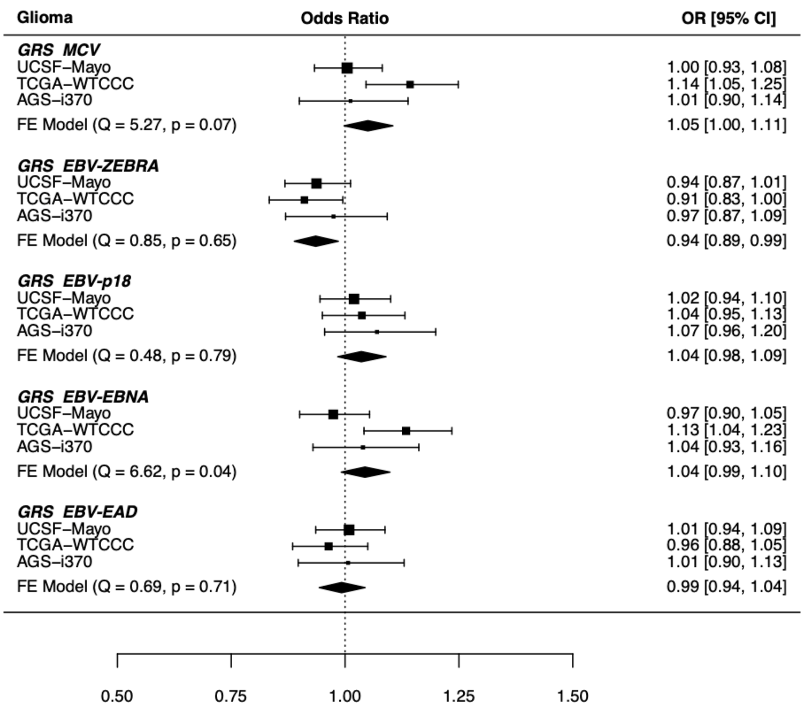


#### S3 B) *IDH* mutated

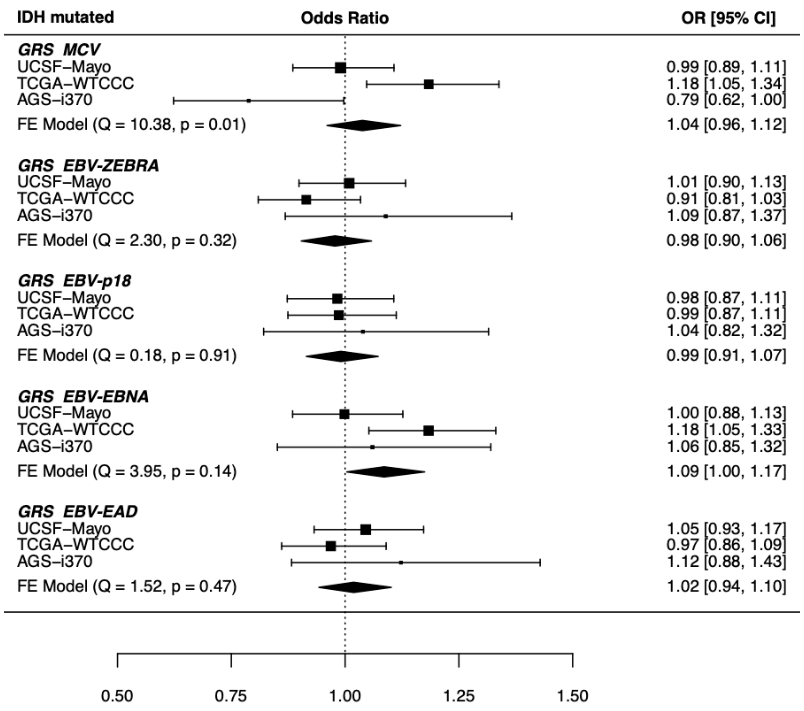

#### S3 C) *IDH* mutated 1p/19q codeleted

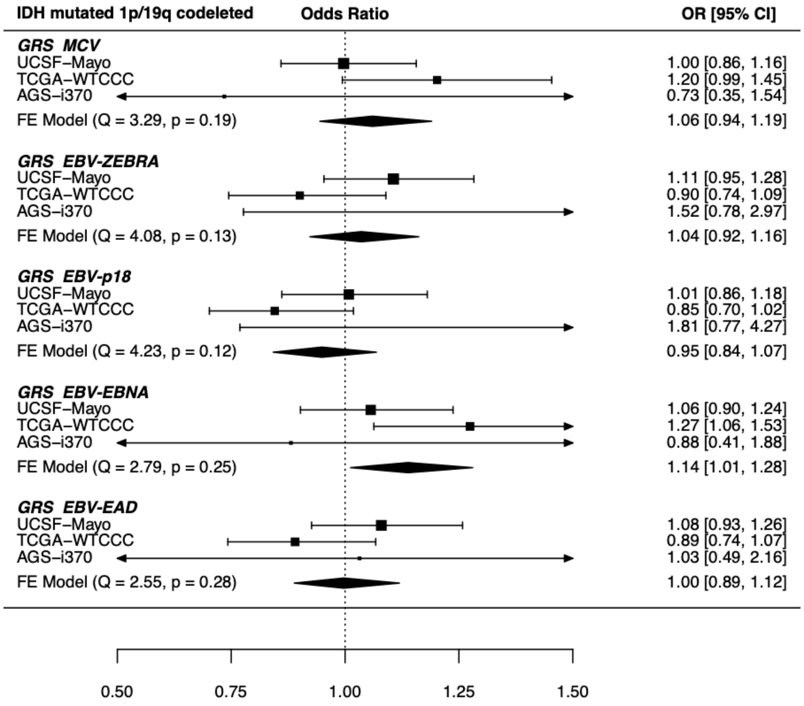


#### S3 D) *IDH* mutated 1p/19q non-codeleted

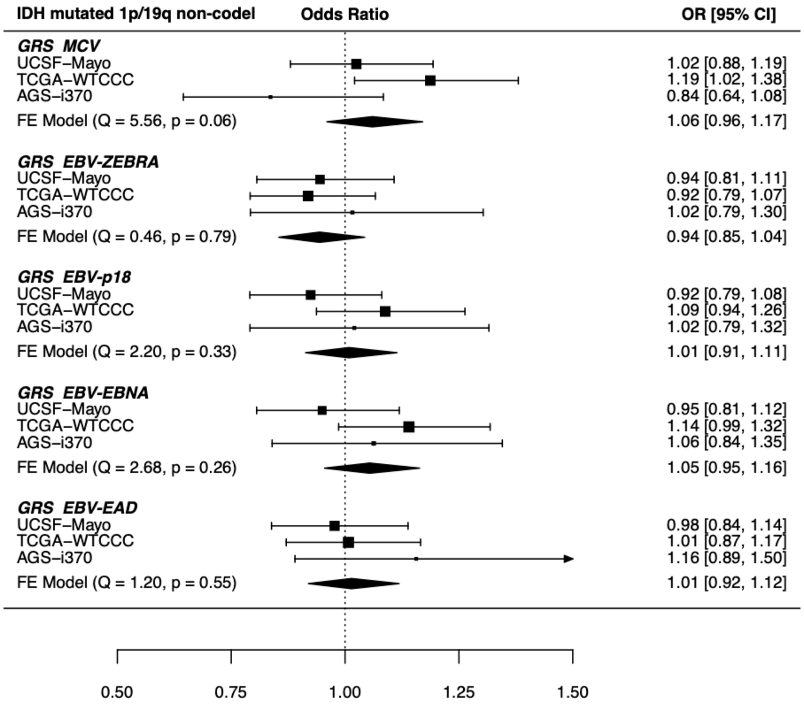

#### S3 E) *IDH* wild type

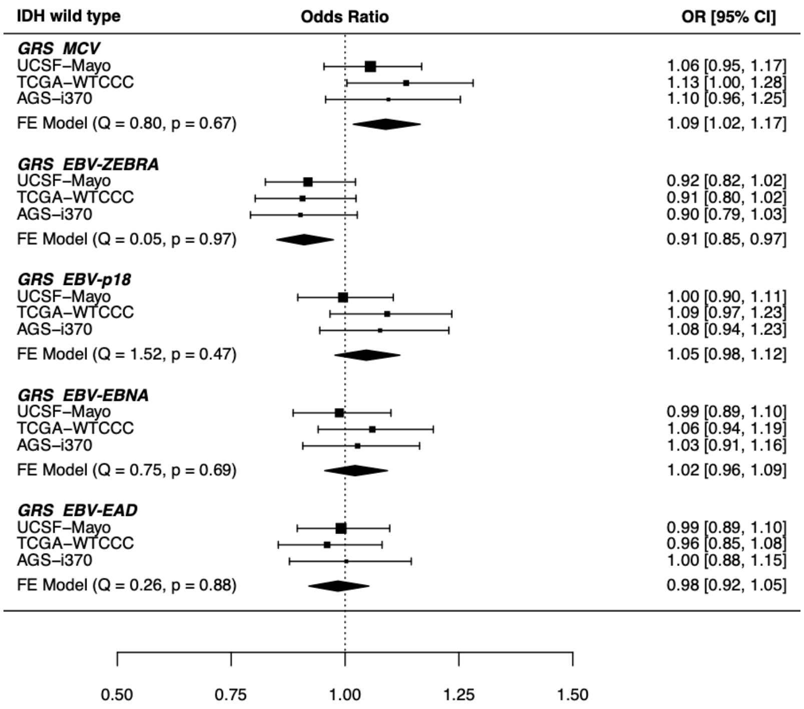


#### S3 F) *IDH* wild type 1p/19q non-codeleted

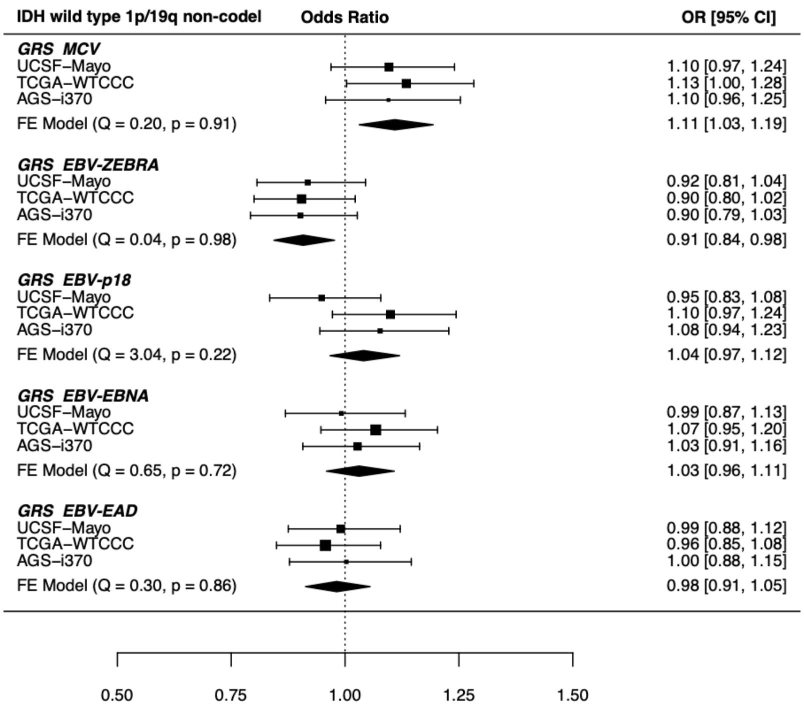

A-F) Forest plot meta-analysis results of all tested GRS-glioma risk associations for the specific primary subtype. Results are reported as odds ratios, along with 95% confidence intervals. Briefly, each header indicates the studied glioma molecular subtype, within are each GRSs associations with the subtype, and the 95% confidence interval of each study-specific effect. The diamond visualizes the 95% confidence interval for the fixed effect (FE) meta-analysis across studies. Each meta-analysis was tested for between-study heterogeneity (Q statistic), with $p<0.05$ indicating evidence of study-specific associations. GRS-subtype tests with evidence of significant heterogeneity were re-analyzed using a random-effects meta-analysis (results not reported). No new suggestive associations were found after re-analysis.

### Figure S4: All GRS-Glioma survival meta result forest plots, by antigen

#### S4 A) Glioma overall

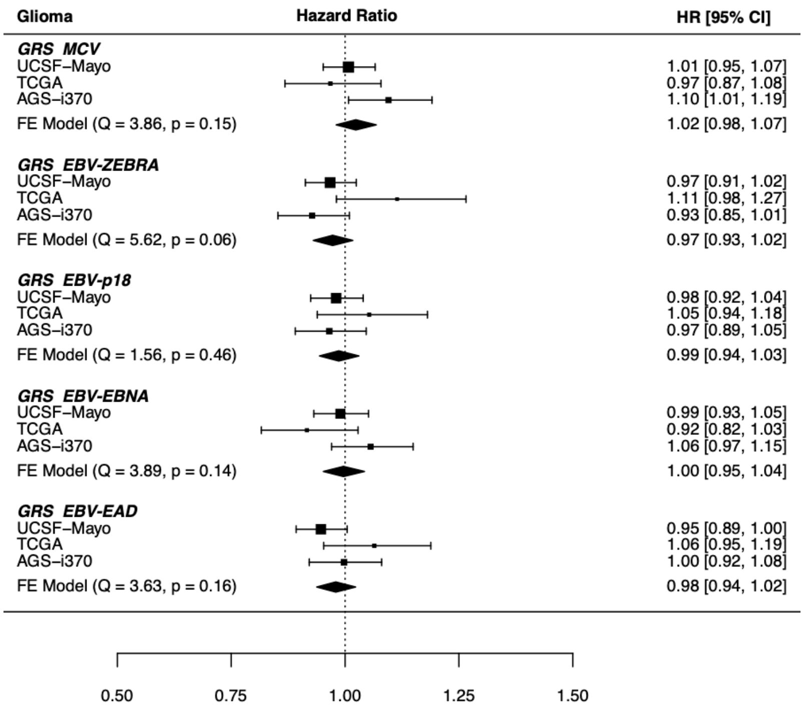


#### S4 B) *IDH* mutated

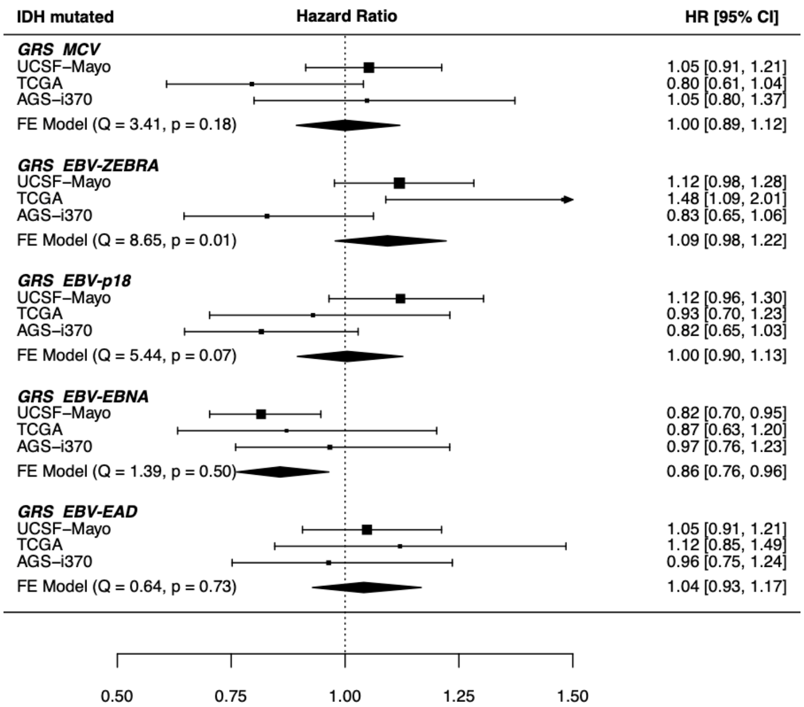

#### S4 C) *IDH* mutated 1p/19q codeleted

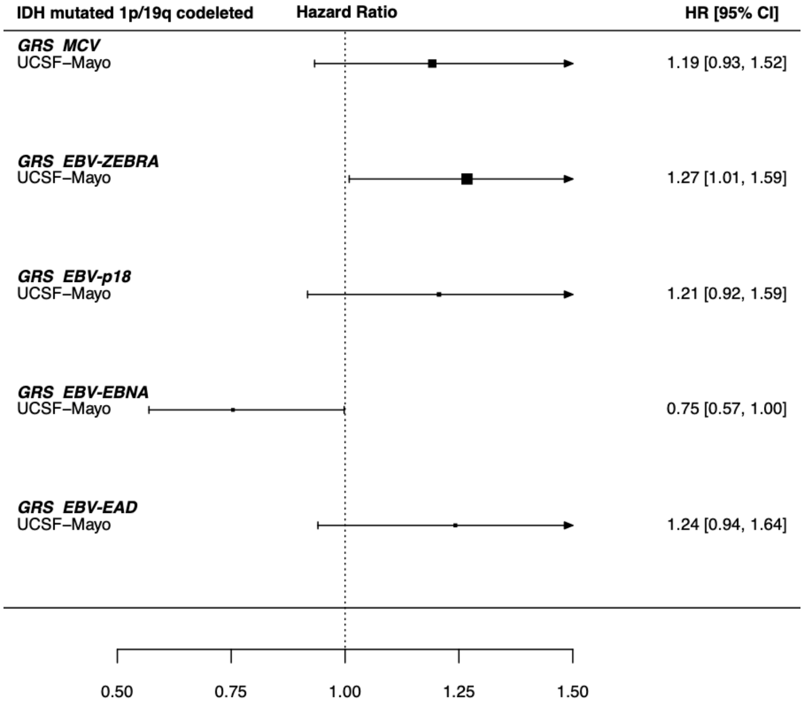


#### S4 D) *IDH* mutated 1p19 non-codeleted

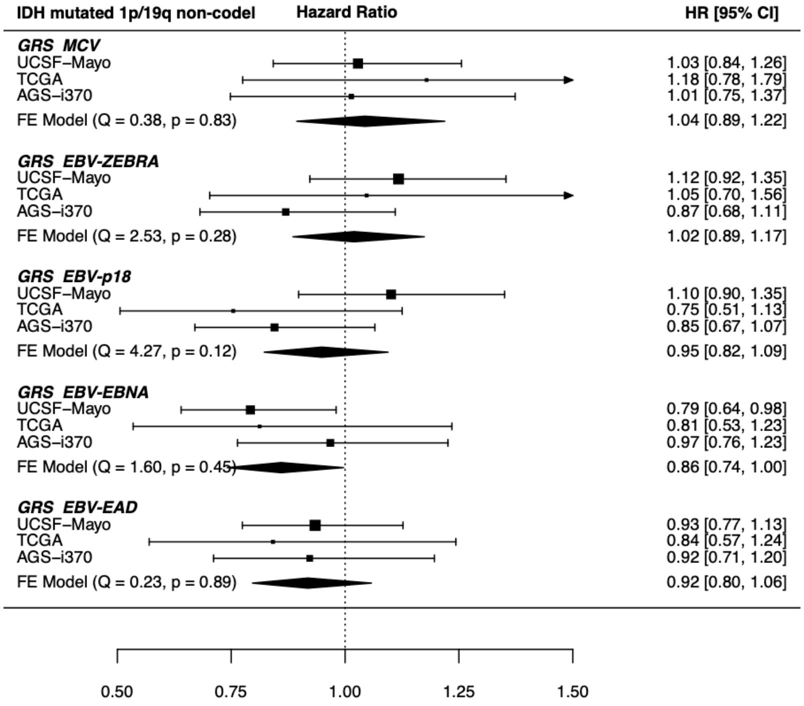

#### S4 E) *IDH* wild type

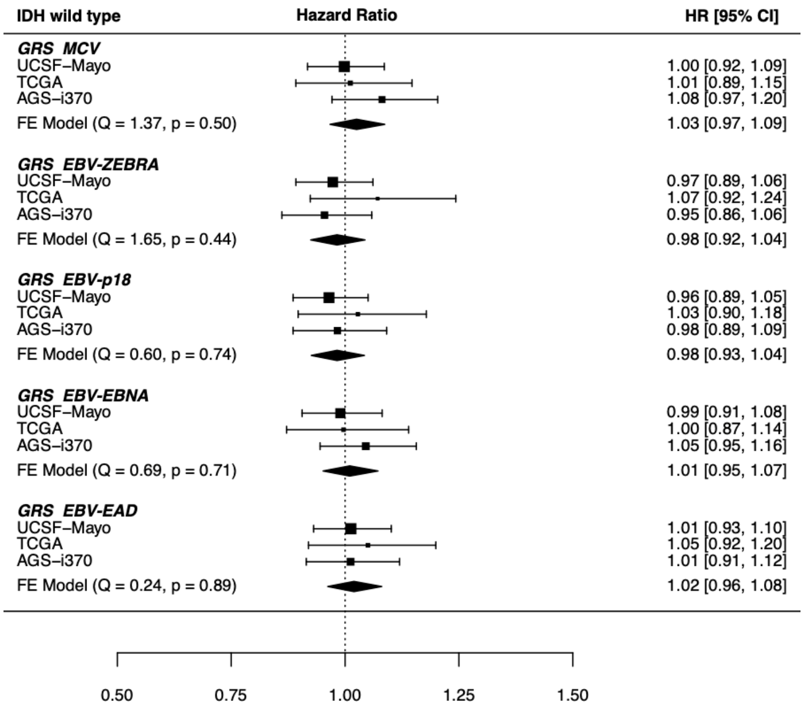


#### S4 F) *IDH* wild type 1p/19q non-codeleted

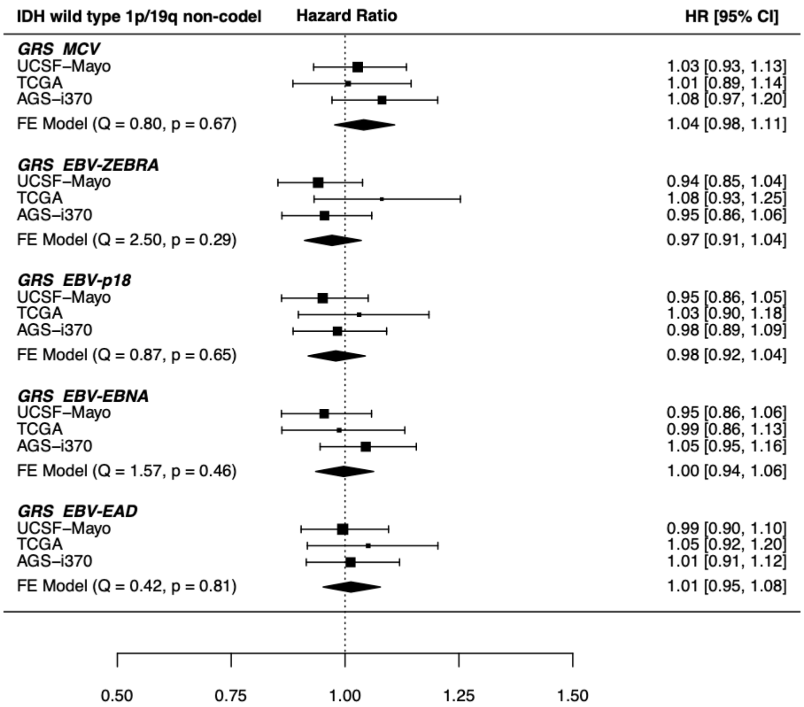

A-F) Forest plot meta-analysis results of all tested GRS-glioma survival associations. Results are reported as hazard ratios, along with 95% confidence intervals. Briefly, each header indicates the studied glioma molecular subtype, within are each GRSs associations with the clinical outcomes of that subtype, and the 95% confidence interval of each study-specific effect. The diamond visualizes the 95% confidence interval for the fixed effect (FE) meta-analysis across included studies. Studies which had an insufficient number of cases/events in a subtype were not included in the meta-analysis and no meta-analysis was performed if only one study was included. Each meta-analysis was tested for between-study heterogeneity (Q statistic), with $p<0.05$ indicating evidence of study-specific associations. GRS-subtype tests with evidence of significant heterogeneity were re-analyzed using a random-effects meta-analysis (results not reported). No new suggestive associations were found after re-analysis.

### Figure S5: Glioma survival Kaplan-Meier curves by antigen GRS and subtype

#### S5 A) Glioma overall

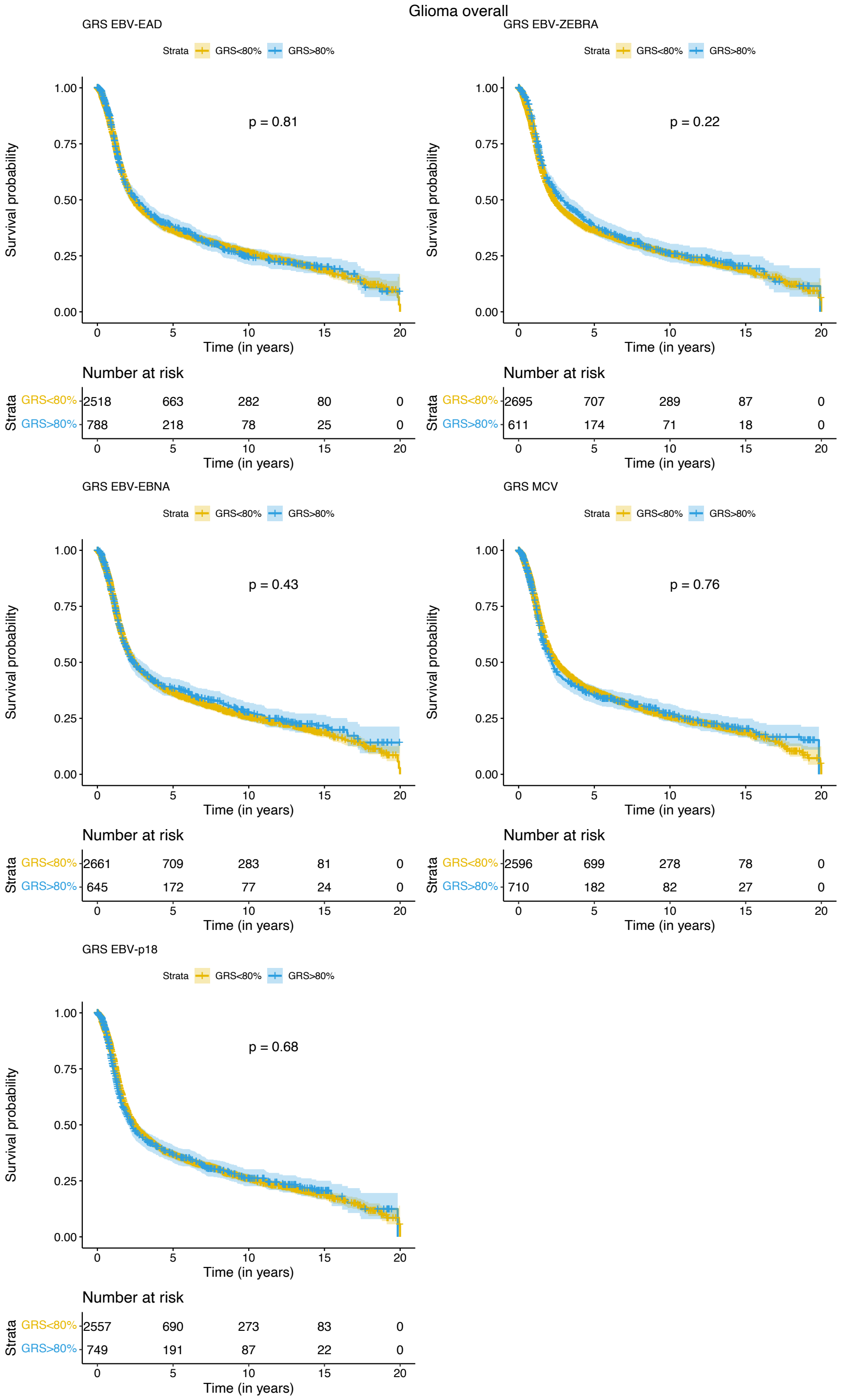

### S5 B) *IDH* mutated

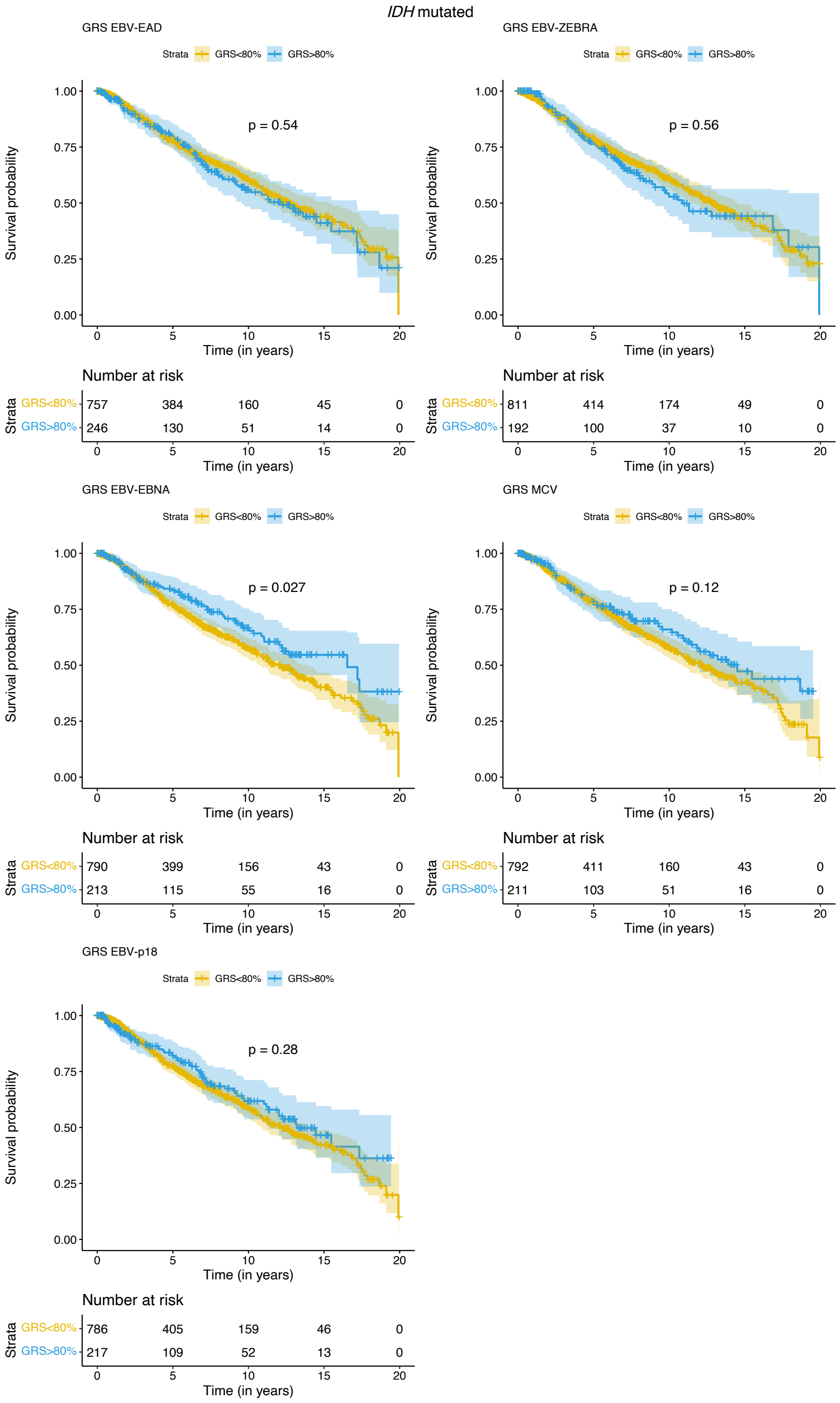

### S5 C) *IDH* mutated 1p/19q codeleted

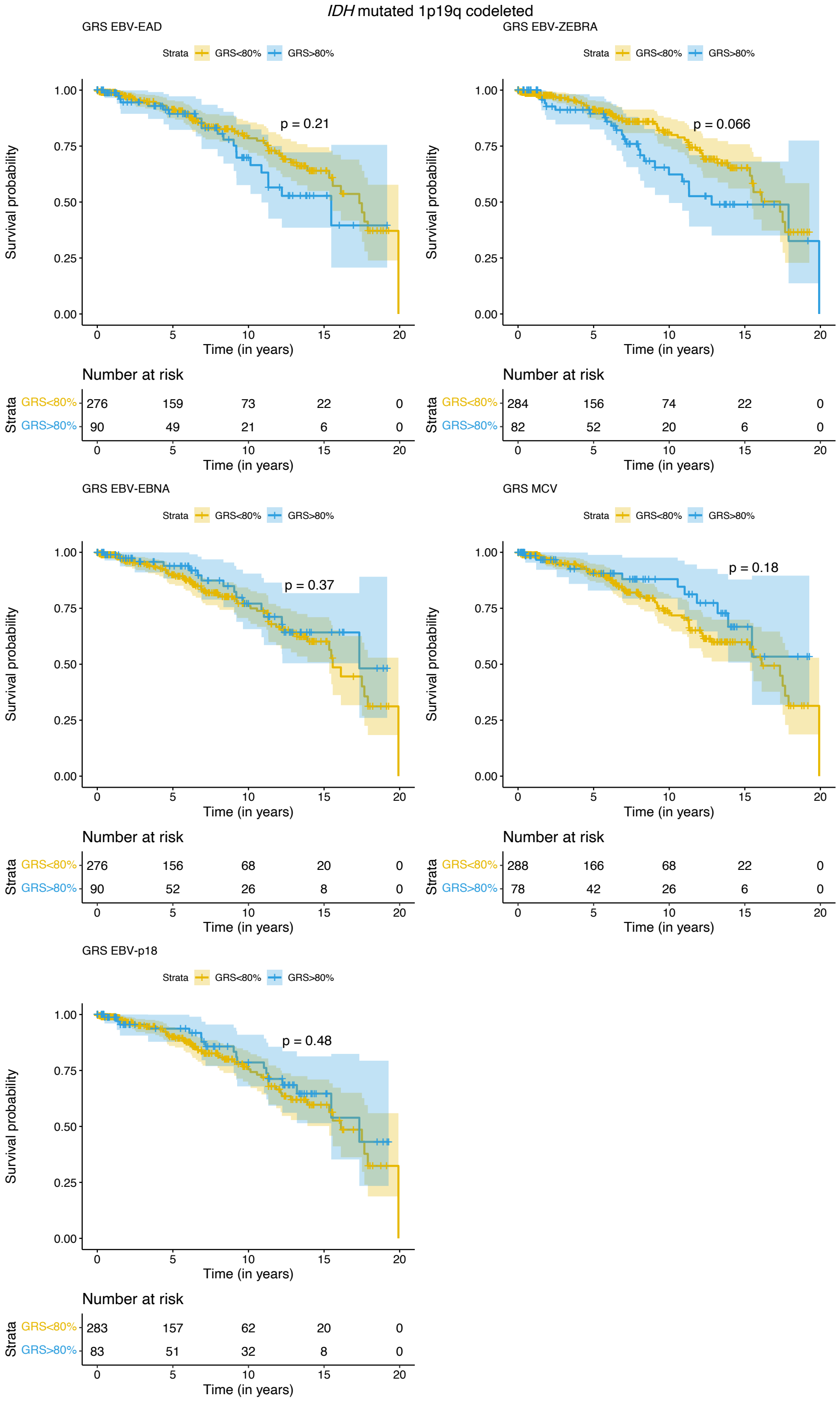

### S5 D) *IDH* mutated 1p19 non-codeleted

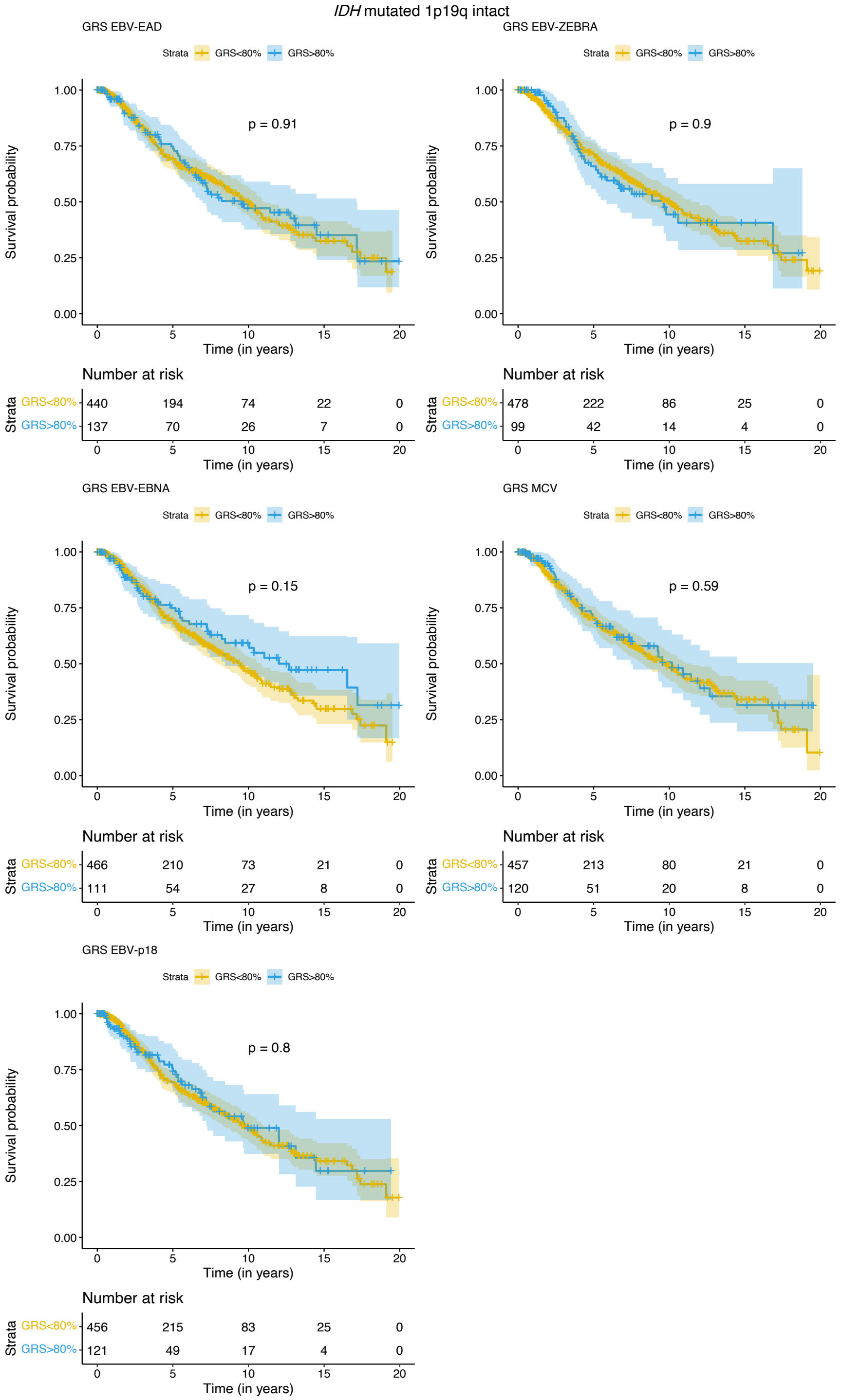

#### S5 E) *IDH* wild type

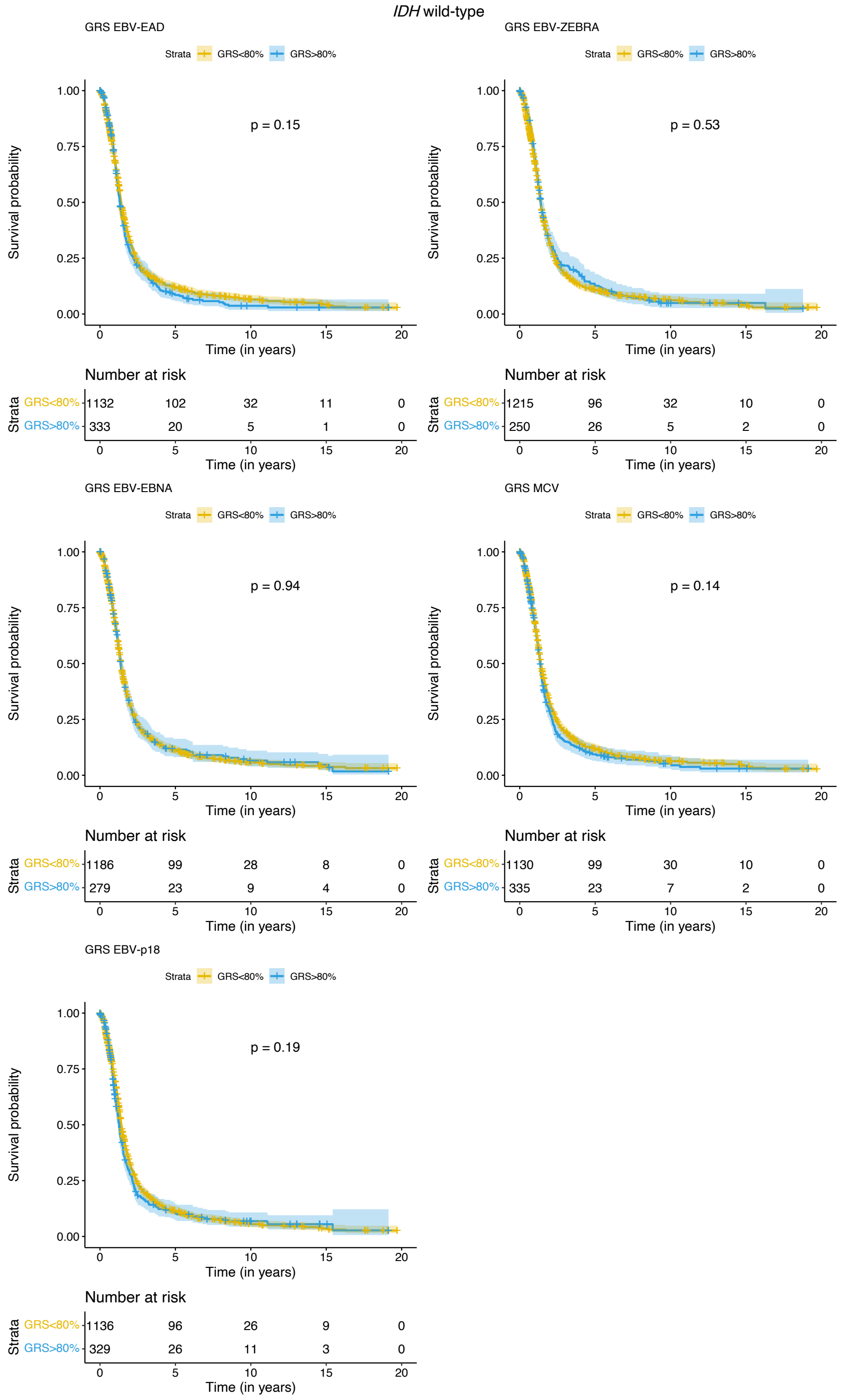

### S5 F) *IDH* wild type 1p/19q non-codeleted

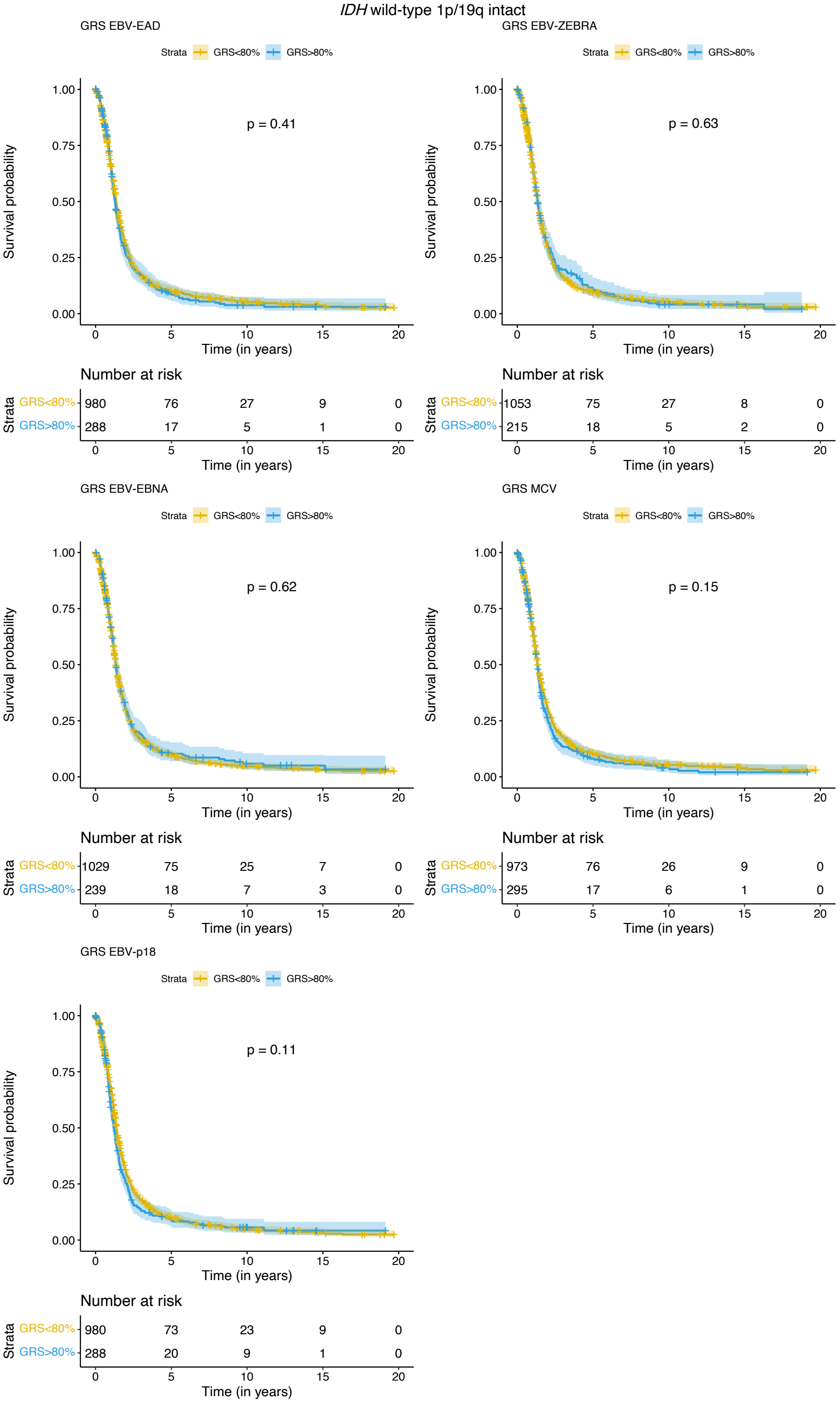

A-F) Kaplan-Meier curves for GRS associations with the survival in the specified molecular glioma subtype. Each plot represents a visualization of the association of a specific viral antigen GRS with clinical outcomes in the molecular subtype. To visualize, each antigen's unnormalized GRS scores across the included studies were binned based on the case-specific 80$^{th}$ percentile score in the UCSF-Mayo dataset. P-values included on each plot are results of a log-rank test for difference between the two curves. Below each set of curves provides the number of cases surviving beyond that time point, for each of the two GRS groups.
